# Disentangling heterogeneity of psychosis expression in the general population: sex-specific moderation effects of environmental risk factors on symptom networks

**DOI:** 10.1101/2021.05.06.21256748

**Authors:** Linda T. Betz, Nora Penzel, Marlene Rosen, Kamaldeep Bhui, Rachel Upthegrove, Joseph Kambeitz

**Author notes:** **Corresponding Author:** Linda T. Betz, Department of Psychiatry and Psychotherapy, Faculty of Medicine and University Hospital of Cologne, Kerpener Str. 62, 50937 Cologne.

## Abstract

**Background:** Psychosis expression in the general population may reflect a behavioral manifestation of the risk for psychotic disorder. It can be conceptualized as an interconnected system of psychotic and affective experiences; a so-called ‘symptom network’. Differences in demographics, as well as exposure to adversities and risk factors, may produce substantial heterogeneity in symptom networks, highlighting potential etiological divergence in psychosis risk.

**Methods:** To explore this idea in a data-driven way, we employed a novel recursive partitioning approach in the 2007 English National Survey of Psychiatric Morbidity survey (*N* = 7,242). We sought to identify ‘network phenotypes’ by explaining heterogeneity in symptom networks through potential moderators, including age, sex, ethnicity, deprivation, childhood abuse, separation from parents, bullying, domestic violence, cannabis use, and alcohol.

**Results:** Sex was the primary source of heterogeneity in symptom networks. Additional heterogeneity was explained by interpersonal trauma (*childhood abuse, domestic violence*) in women and *domestic violence, cannabis use, ethnicity* in men. Among women, especially those exposed to early interpersonal trauma, an affective loading within psychosis may have distinct relevance. Men, particularly those from minority ethnic groups, demonstrated a strong network connection between hallucinatory experiences and persecutory ideation.

**Conclusion:** Symptom networks of psychosis expression in the general population are highly heterogeneous. The structure of symptom networks seems to reflect distinct sex-related adversities, etiologies, and mechanisms of symptom-expression. Disentangling the complex interplay of sex, minority ethnic group status, and other risk factors may help optimize early intervention and prevention strategies in psychosis.

## Introduction

Recent research has advanced our understanding of psychosis through so-called ‘symptom networks’, i.e., causal systems of individual interacting experiences and symptoms (Betz et al., 2020; Hardy, O’Driscoll, Steel, van der Gaag, & van den Berg, 2020; Isvoranu, Borsboom, van Os, & Guloksuz, 2016; Isvoranu et al., 2019, 2017; Moffa et al., 2017; Murphy, McBride, Fried, & Shevlin, 2018; Robinaugh, Hoekstra, Toner, & Borsboom, 2020). Complex interactions between specific psychotic as well as non-psychotic experiences (e.g., depression, anxiety) in the general population may predate onset of psychosis in clinical settings (Guloksuz et al., 2016, 2015; Kelleher et al., 2012; Linscott & van Os, 2013; Murphy et al., 2018; van Os & Reininghaus, 2016). Additional lines of evidence indicate that there is considerable etiological continuity between subclinical and clinical levels of psychosis (Binbay et al., 2012; DeRosse & Karlsgodt, 2015; Kelleher & Cannon, 2011; Linscott & van Os, 2013). Thus, examining the symptom network structure of a transdiagnostic psychosis phenotype, reflecting a behavioral manifestation of risk for psychotic disorder in the general population that blends gradually into clinical syndromes, may help to better understand etiological mechanisms in psychosis and to develop prevention strategies (Bebbington, 2015; Binbay et al., 2012; DeRosse & Karlsgodt, 2015; Isvoranu et al., 2016; Kelleher & Cannon, 2011; Linscott & van Os, 2013; Robinaugh et al., 2020; van Os & Reininghaus, 2016).

Importantly, symptomatology and involved etiological mechanisms in psychosis expression are highly variable by specific at risk groups (Bentall, Wickham, Shevlin, & Varese, 2012; Isvoranu et al., 2016; Linscott & van Os, 2013; van Os & Reininghaus, 2016). For example, in line with the theory of an affective pathway to psychosis, early traumatic events are strongly associated with connections between affective and psychotic symptomatology (Myin-Germeys & van Os, 2007; Upthegrove et al., 2015; van Nierop et al., 2015). In the presence of heterogeneity, averaged network models of psychosis may obscure important distinctions in relevant etiological mechanisms across specific risk groups (Jones, Mair, Simon, & Zeileis, 2020; Moriarity, van Borkulo, & Alloy, 2020). Thus far, however, heterogeneity in symptom networks of psychosis has been either overlooked or addressed in a partial way on a single candidate risk factor (such as sex, cannabis use or socioeconomic background) at specific thresholds, or using summed environmental risk scores (Betz et al., 2020; Guloksuz et al., 2016; Isvoranu et al., 2016; Wüsten et al., 2018), which lose specificity and relevance for real work prevention and intervention.

The characterization of ‘network phenotypes’ based on a comprehensive set of environmental and demographic factors may explain heterogeneity; that is the structure of symptomatology is a function of types, combinations, and intensity of etiological loads in psychosis expression (Jones et al., 2020; Moriarity et al., 2020). With the goal of characterization of network phenotypes in mind, the current study uses novel work on recursive partitioning, a data-driven, explorative statistical technique that can sequentially extract isolated and combined moderation effects of a large set of environmental and demographic factors on symptom networks, without a priori specification of thresholds or combinations of risk factors (Jones et al., 2020; Strobl, Malley, & Tutz, 2009). Recursive partitioning identifies network phenotypes that are maximally distinct from each other (Jones et al., 2020; Zeileis, Hothorn, & Hornik, 2008).

We used recursive partitioning to define meaningful network phenotypes of psychosis expression in the general population, using the 2007 Adult Psychiatric Morbidity in England Survey (APMS; (National Centre for Social Research, University of Leicester, 2017). We hypothesized that exposure to environmental risk, if identified as defining a network phenotype, would be characteristically associated with more densely connected symptom networks when compared with samples not exposed to that specific environmental risk (Guloksuz et al., 2016, 2015; Isvoranu et al., 2016; Lin, Fried, & Eaton, 2019; Russell, Keding, He, Li, & Herringa, 2020). We also aimed to test whether the strength of connections between individual symptoms differed between network phenotypes.

## Method

### Data analytic strategy

We conducted all analyses in the *R* language for statistical computing, version 4.0.4. Throughout, we considered a significance level of α = .05. Data of the 2007 APMS (National Centre for Social Research, University of Leicester, 2017) used in the analyses are available from the UK Data Service (https://ukdataservice.ac.uk/). Code to reproduce the analyses can be accessed at www.github.com/LindaBetz/APMS_NetworkTree.

### Sample

We present analyses based on the 2007 APMS of adults living in private households aged 16 and above who were recruited using a stratified multistage random probability sampling strategy (*N* = 7,403) (McManus, Meltzer, Brugha, Bebbington, & Jenkins, 2009; Singleton, Bumpstead, O’Brien, Lee, & Meltzer, 2003). Methods, procedures, and full details on sample characteristics have been described previously (McManus et al., 2009). For the present analyses, we excluded participants with missing values in the variables of interest, given that the methods employed do not allow missings. For comparing sample characteristics of included and excluded participants, we used permutation tests as implemented in the *R* package ‘coin’ (Hothorn, Hornik, van de Wiel, & Zeileis, 2008).

### Assessment of symptomatology

Selection and definition of symptom variables followed a previously published network analysis using data from the 2007 APMS (Moffa et al., 2017), including measures from an affective domain (worry, sleep disturbance, generalized anxiety, and depression), and from a psychotic domain (persecutory ideation and hallucinatory experiences). All symptom variables in the network were coded in binary form (present or absent). For details on these assessments, see supplementary method 1.

### Assessment of environmental and demographic risk factors

Environmental risk factors comprised psychosocial adversities in the form of physical abuse and sexual abuse before the age of 16, separation from parents until the age of 16 (local authority care and/or institutional care), lifetime experiences of bullying, and lifetime experiences of domestic violence. Additionally, we included sex, age, ethnic origin (White, Black, South Asian, Mixed/Other), cannabis use in the past year, alcohol use, and socioeconomic deprivation. For details on these assessments, see supplementary method 2.

### Identification of network subgroups via recursive partitioning

In a first step, we estimated a partial correlation network (without regularization) based on the full sample, using the *R* package ‘qgraph’, version 1.6.5 (Epskamp, Cramer, Waldorp, Schmittmann, & Borsboom, 2012). A partial correlation network depicts unique pairwise associations between variables (‘edges’ in network terminology), i.e., the share of the association between two variables that remains after controlling for all other variables in the network (Epskamp, Borsboom, & Fried, 2018). We estimated the underlying zero-order correlations between the binary items using Pearson’s φ, as recommended when employing recursive partitioning on binary data (Jones et al., 2020). The stronger the partial correlation between two variables, the more likely it is that they co-occur, controlling for the other variables under consideration.

Second, we used a model-based recursive partitioning approach to identify meaningful subgroups of symptom networks given the included environmental and demographic factors, as implemented in the *R* package ‘networktree’, version 1.0.1 (Jones et al., 2020). In brief, recursive partitioning sequentially creates a decision tree by either splitting or not splitting the sample along a set of potential moderating variables (Strobl et al., 2009; Zeileis et al., 2008). The ‘networktree’ approach (figure 1) determines sample splits based on significant invariance in the correlation matrix of the network variables under consideration, yielding non-overlapping partitions of the sample with maximally heterogeneous symptom networks (Jones et al., 2020). For a detailed account, we refer to supplementary method 3 and available methodological articles (Jones et al., 2020; Strobl et al., 2009; Zeileis et al., 2008). For plotting, we transformed the correlation matrices to partial correlation matrices using the *R* package ‘qgraph’, such that edges reflect unique associations between two variables.

**Figure 1.**
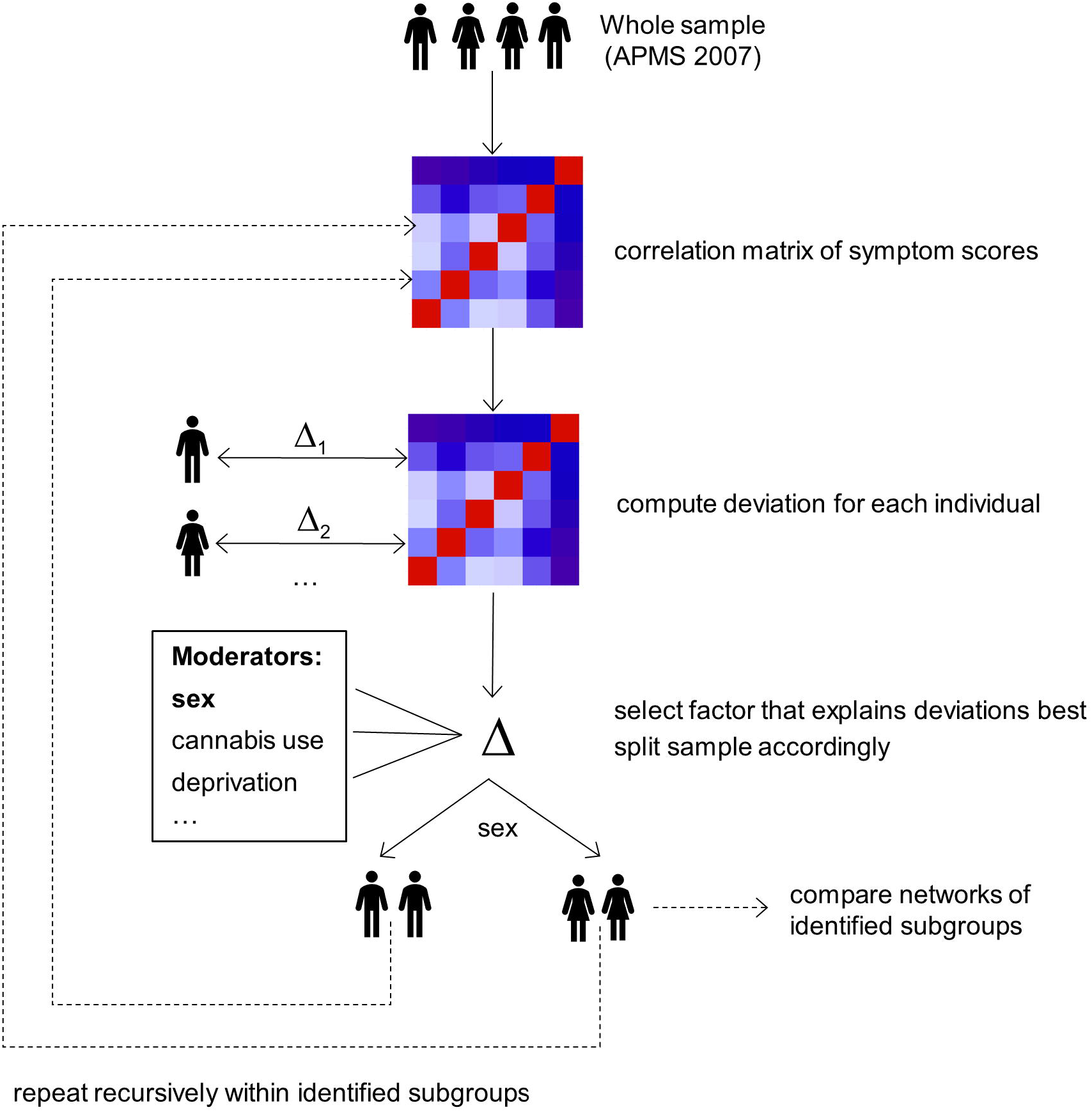
Recursive partitioning for symptom networks as applied to data from the 2007 Adult Psychiatric Morbidity Survey (APMS) study. The goal is to assess which of the included demographic and risk factors capture individual deviations from the correlation matrix of symptom scores, which underlies symptom networks. Starting with the whole sample, individual deviations from the correlation matrix of symptom scores are computed via a log-likelihood-based score function. The variable that explains these deviations best, as determined by a minimum *p*-value strategy at Bonferroni-corrected α, is selected (here: sex), and the sample split accordingly. Within the identified subgroups, the procedure is repeated recursively until no significant deviations, i.e., heterogeneity, is detected. We compared symptom networks of the identified subgroups in terms of global strength and individual edge weights. For a detailed account of the method, see supplementary method 3 and Jones et al. (2020).

### Comparison of identified subgroups

To delineate specific network differences between the identified subgroups (i.e., differences between subgroups as defined by a splitting factor in the recursive partitioning approach), we compared the overall strength of symptom connections, defined as the absolute sum of all individual partial correlation coefficients in the network (global strength; S), and differences in estimates of individual partial correlation coefficients (individual edge weights; ρ) within a Bayesian framework, using the *R* package ‘BGGM’, version 2.0.2 (Williams, 2021; Williams & Mulder, 2019; Williams, Rast, Pericchi, & Mulder, 2020). Specifically, we used posterior predictive checks for assessing differences in overall connection strength (Williams et al., 2020), and evaluated the posterior distribution for each difference in partial correlation coefficients, where we deemed a difference significant if the 95% credible interval did not contain 0 (Williams, 2021)*.P*-values derived from recursive partitioning are denoted as *p*_RP_, while *p*-values derived from post-hoc comparisons implemented in the package ‘BGGM’ are denoted as *p*_BGGM_.

### Robustness Analyses

We used the *R* package ‘bootnet’, version 1.4.3 (Epskamp et al., 2018) to conduct robustness analyses to check stability and accuracy of the results. We investigated stability of symptom networks estimated in the full sample and identified subgroups by testing sensitivity to dropping cases. Specifically, we assessed the degree to which edge weights remained the same after re-estimating the networks with less cases via the correlation stability (CS) coefficient. The CS coefficient represents the maximum proportion of cases that can be dropped, such that the correlation between original edge weights and edge weights of networks based on subsets is 0.7 or higher (95% confidence). The CS-coefficient should preferably be above 0.5 (good stability), and not be below 0.25 (acceptable stability) (Epskamp et al., 2018). To investigate the accuracy of individual edge weights estimates across the networks in the full sample and identified subgroups, participants were randomly resampled 5000 times, and the bootstrapped confidence intervals (CIs) of the edge weights were estimated.

## Results

### Sample

Following removal of 161 participants (2.2% of the whole sample) with missing values in the variables of interest, the final sample comprised 7,242 participants, 56.8% of whom were women, with an average age of 50 (*IQR* = 30) years. Participants excluded due to missing data were on average older, less White and reported lower proportions of alcohol use and hallucinatory experiences, and higher proportions of depressive symptoms (supplementary table 1).

### Network variables and potential moderators

Table 1 presents positive endorsement and characteristics of the network variables and potential moderating risk factors in the sample. The most prevalent symptom was worry, and the most prevalent risk factor bullying.

### Overall symptom network structure and subgroups

The partial correlation network estimated in the full sample suggested positive relationships between all symptoms, with a mean edge weight of 0.11. Partial correlations within each symptom domain were, on average, stronger than between the domains. Recursive partitioning revealed that six of the tested demographic and environmental risk factors were linked to significant heterogeneity in symptom networks and split the sample accordingly in a hierarchical fashion: *sex, childhood sexual abuse, childhood physical abuse, domestic violence, cannabis use, ethnicity*. Partial correlation matrices for the plotted networks are available at the linked GitHub repository. Sex was the primary source of heterogeneity (figure 2a, *p*_RP_ < .001): the network of women was overall significantly less strongly connected (ΔS = −0.17, *p*_BGGM_ = .002), and featured a significantly stronger connection between depression and hallucination (Δρ = 0.06), and a significantly weaker connection between sleep problems and persecutory ideation (Δρ = −0.07) than the network of men. This means that in women, depression and hallucination were more likely to co-occur than in men, while sleep problems and persecutory ideation were less likely to co-occur than in men. For networks of women and men, see figure 3.

**Figure 2.**
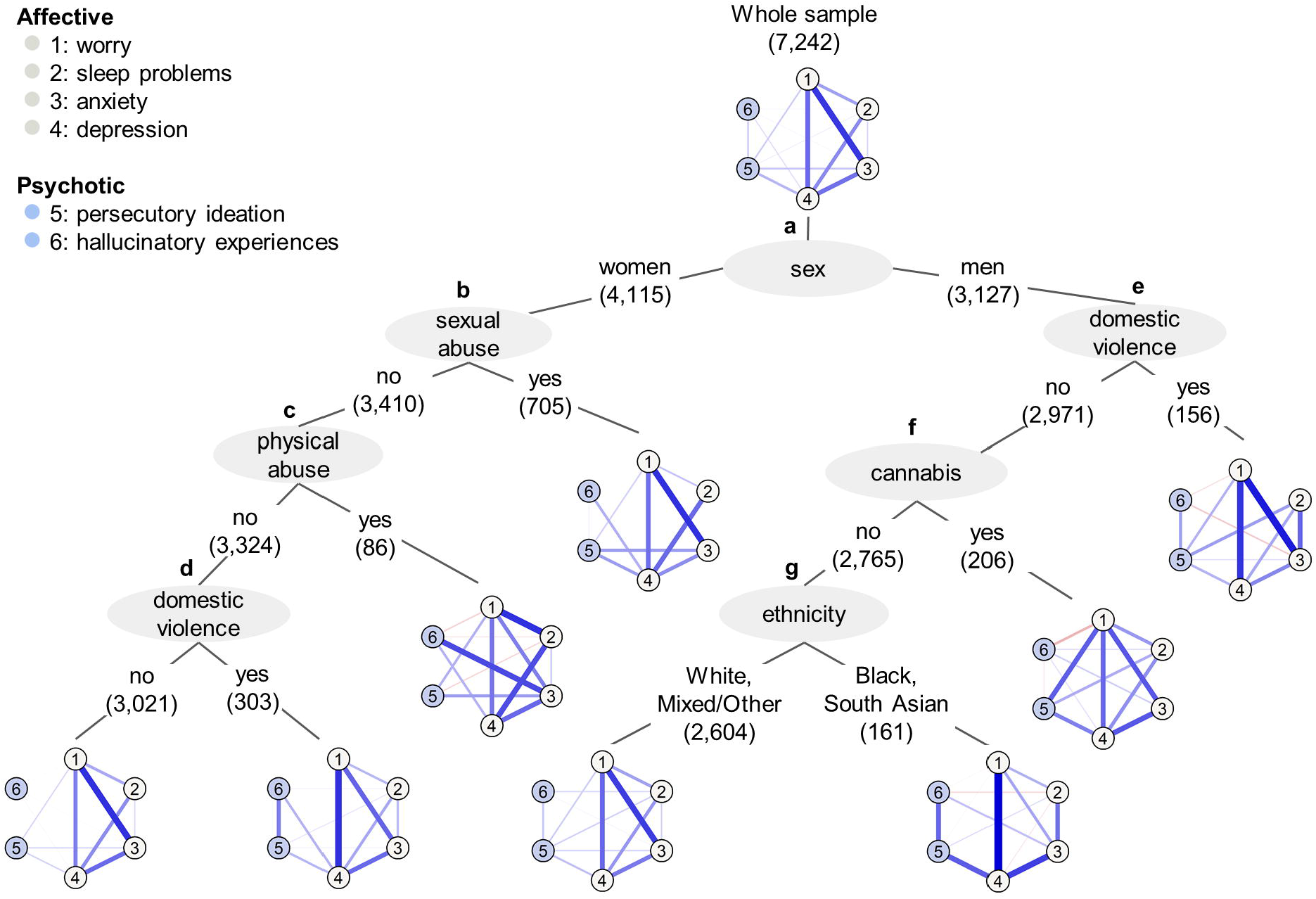
Results from recursive partitioning, depicted as a decision tree of partial correlation networks. Numbers behind splitting factors give the sample size retained after the corresponding sample split. Symptom domains are differentiated by color. The thicker and less transparent the edge, the stronger the partial correlation between two symptoms. Blue (red) edges indicate positive (negative) relationships. To ensure visual comparability, edge weights were scaled identically across all networks. Only connections representing edge weights larger than 0.01 are depicted.

**Figure 3.**
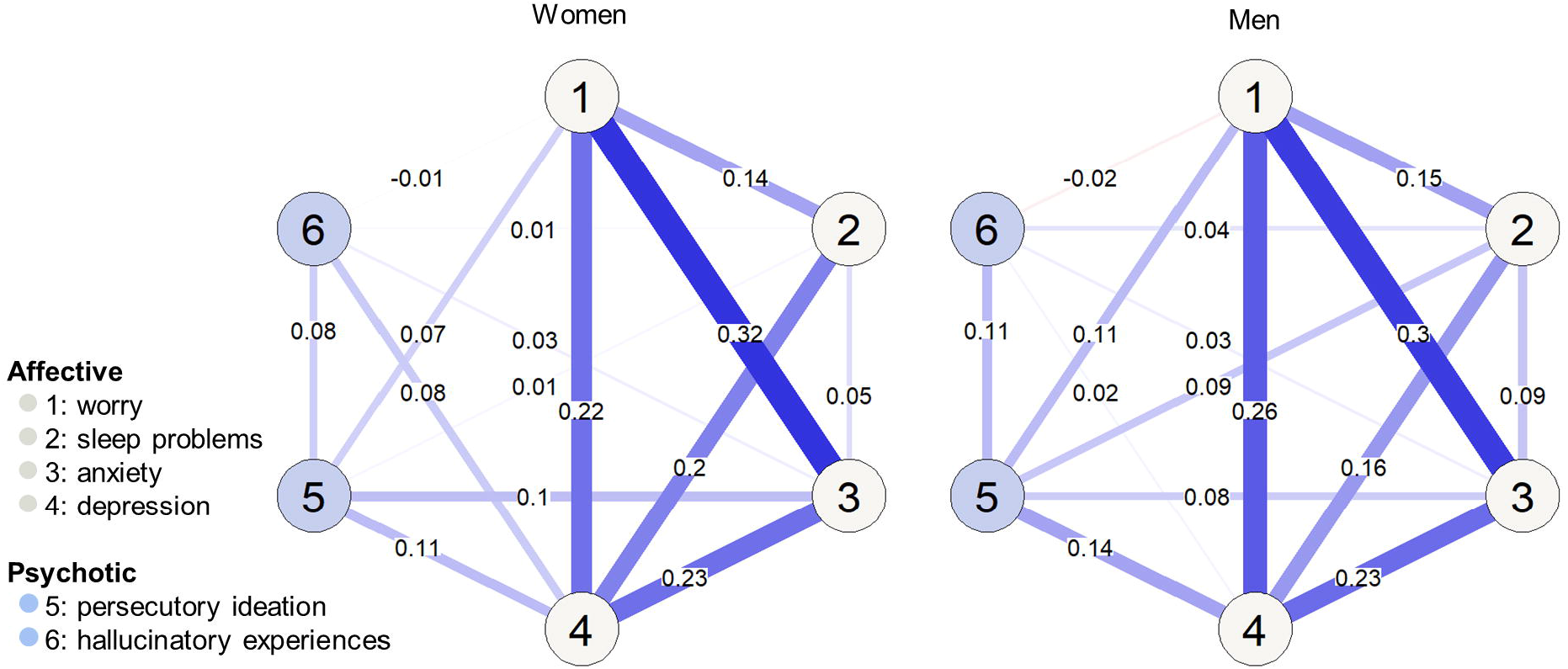
Partial correlation networks estimated in women (*n* = 4,115) and men (*n* = 3,127). Sex was identified as the first split in the recursive partitioning approach, suggesting that sex was the primary source of heterogeneity in symptom networks. Symptom domains are differentiated by color. The thicker and less transparent the edge, the stronger the partial correlation between two symptoms. Blue (red) edges indicate positive (negative) relationships. To ensure visual comparability, edge weights were scaled identically across both networks.

Distinct risk factors explained further heterogeneity in symptom networks of women and men, yielding 8 different network phenotypes in total. Among women, experiences of childhood sexual abuse were the major source of heterogeneity in symptom networks (figure 2b, *p*_RP_ = .016) linked to a stronger connection between anxiety and persecutory ideation (Δρ = 0.09). The difference in global strength of the symptom networks of women who reported sexual abuse and those who did not was not significant (ΔS = 0.08, *p*_BGGM_ = .482). Among women who reported no childhood sexual abuse, exposure to childhood physical abuse explained further heterogeneity (figure 2c, *p*_RP_ = .015), and was associated with a significantly stronger association between anxiety and hallucinations (Δρ = 0.26). Corresponding symptom networks did not differ significantly in global strength (ΔS = 0.70, *p*_BGGM_ = .204). Finally, among those women that reported neither sexual nor physical abuse, exposure to domestic violence (figure 2d, *p*_RP_ = .012) was linked to a stronger connection between worry and depression (Δρ = 0.12), as well as persecutory ideation and hallucinations (Δρ = 0.19). The difference in global strength of the corresponding symptom networks was not significant (ΔS = 0.32, *p*_BGGM_ = .113).

Among men, in those who reported having experienced domestic violence (figure 2e, *p*_RP_ = .007) the connection between sleep problems and anxiety was significantly stronger than in men who did not report past domestic violence (Δρ = 0.17). Global strength was not significantly different between the corresponding networks (ΔS = 0.26, *p*_BGGM_ = .376). Second, in men not reporting past domestic violence, cannabis use in the past year (figure 2f, *p*_RP_ = .009) was associated with a significantly increased connection between worry and persecutory ideation (Δρ = 0.17), and a significantly weaker connection between hallucination and persecutory ideation (Δρ = −0.18). Global strength of the corresponding symptom networks did not differ significantly (ΔS = 0.33, *p*_BGGM_ = .126). Finally, men reporting neither domestic violence nor cannabis use were further split by ethnic background (figure 2g, *p*_RP_ = .011): the network of men with a Black or South Asian ethnic background was overall significantly more strongly connected (ΔS = 0.76, *p*_BGGM_ = .003), and showed stronger connections between worry and depression (Δρ = 0.25), sleep problems and anxiety (Δρ = 0.20), anxiety and depression (Δρ = 0.16), depression and persecutory ideation (Δρ = 0.21), as well as persecutory ideation and hallucinatory experiences (Δρ = 0.20), and a weaker connection between sleep problems and depression (Δρ = −0.23) than the network of men from a White or Mixed ethnic background.

Age of the respondent, alcohol use, bullying, separation experiences, and socioeconomic deprivation were not identified as relevant sources of heterogeneity in symptom networks. Repeating analyses based on data from women and men separately yielded identical results regarding sex-specific moderators (supplementary figure 1).

### Robustness analyses

The network estimated in the full sample, as well as all identified subgroup networks, showed good stability to dropping cases (supplementary table 2). Accuracy analyses showed some relatively wide bootstrapped CIs in some of the identified subgroups with smaller sample sizes. In these cases, we recommend caution when interpreting the strength of weaker edges. Still, the bootstrap mean was generally very close to the sample mean, indicating interpretable results (supplementary figures 2-16).

## Discussion

In the present study, we employed a novel, data-driven recursive partitioning approach in a large national household survey to identify networks of psychotic and affective experiences in the population. Our findings point to considerable heterogeneity, which we explain with several phenotypic systems: six (out of eleven) demographic and environmental risk factors yielded eight different network phenotypes, with sex being the primary source of heterogeneity in symptom networks. Among women and men, different risk factors were related to heterogeneity in symptom networks, suggesting potentially distinct relevance and mechanisms of these risk factors across the sexes, in line with a multidimensional model of sexual differentiation in psychosis risk (Riecher-Rössler, Butler, & Kulkarni, 2018). Overall, our findings on sex and other environmental differences illustrate that the multifactorial and heterogeneous nature of psychosis expression (Isvoranu et al., 2016; Linscott & van Os, 2013; van Os & Reininghaus, 2016) appears to be reflected in symptom networks that differed considerably depending on the type, combination and strength of demographic and environmental risk in a large general population sample.

### Differences in symptom networks of women and men

The identification of multiple network phenotypes substantiates the notion that averaged symptom network models are likely not representative of psychosis expression in the general population (Jones et al., 2020). Rather, observed differences in strength of overall and specific symptom connections may point to diverse etiological mechanisms operating across different demographic and environmental risk factors. Corroborating a growing recognition that understanding variability by sex is central for the development of comprehensive etiological models of psychopathology (Hartung & Lefler, 2019; Hodes & Epperson, 2019; Riecher-Rössler et al., 2018; Rosen, Haidl, Ruhrmann, Vogeley, & Schultze-Lutter, 2019), the primary source of heterogeneity in symptom networks of psychosis was sex.

Specifically, our results highlight how associations between affective and psychotic experiences may be differentially expressed in women and men. Prior research indicates that, following the theory of an affective pathway to psychosis, affective alterations, in particular depression and anxiety, may be fundamental driving forces of psychotic experiences (Betz et al., 2020; Isvoranu et al., 2017; Myin-Germeys & van Os, 2007; Upthegrove et al., 2020; Upthegrove, Marwaha, & Birchwood, 2017; van Nierop et al., 2018). Present findings suggest a particularly strong association between depression and hallucinatory experiences in the network of women compared to men, corroborating the idea that such an affective pathway to psychosis involving depression may be expressed to a greater degree in women, potentially funneled by increased emotional reactivity to life events and daily hassles (Davis, Matthews, & Twamley, 1999; Hodes & Epperson, 2019; Myin-Germeys & van Os, 2007; Stainton et al., 2021). In the symptom network of men, by contrast, a previously identified link between sleep problems and persecutory ideation (Freeman et al., 2010) was stronger, and therefore, possibly more relevant, than in women. An intriguing potential clinical implication to be tested is that men may, on average, profit in particular from the use of interventions for sleep problems with demonstrated benefit for reducing persecutory ideation (Freeman et al., 2017).

### Risk factors explaining heterogeneity in symptom networks of women and men

Among women, heterogeneity in symptom networks of psychosis expression was explained by exposure to interpersonal trauma, including childhood abuse and domestic violence. Specifically, exposure to childhood abuse was linked to stronger associations between anxiety and psychotic experiences. These findings are consistent with previous reports of increased proportions of mixed symptom expression following childhood trauma (Guloksuz et al., 2015; Russell et al., 2020; Upthegrove et al., 2015; van Nierop et al., 2015), but extend the literature by highlighting how sex may be an important determinant in this relationship. Following trauma, women are more likely to blame themselves, to view the world as dangerous, and to hold more negative views of themselves (Davis et al., 1999; Tolin & Foa, 2002). This may facilitate a pathway from distressing interpretations of everyday events, including the experience of anxiety, to threat beliefs feeding into the formation of psychotic experiences, as proposed in cognitive models of psychosis (Freeman, 2007; Garety, Kuipers, Fowler, Freeman, & Bebbington, 2001; Hardy et al., 2020). Overall, the idea that a pathway from anxiety to psychotic experiences may be particularly relevant among women with a history of childhood abuse has potentially important repercussions for clinical practice and deserves further investigation (Bloomfield et al., 2020). Moreover, at a population level, it may well be that links between affective and psychotic experiences following childhood abuse are manifestations of personality function. The interplay between borderline personality functioning and affective instability, also involving psychosis, and subclinical and clinical levels of psychosis warrant further investigations (Barnow et al., 2010).

Among men, cannabis use and minority ethnic group status were identified as potential sources of heterogeneity in network connections between psychotic and affective symptoms. Most striking differences were evident in the symptom network of men with a minority ethnic group status reporting no domestic violence or cannabis use. Documented variations in experience and reporting of hallucinations (Vanheusden et al., 2008) and delusions (Berg et al., 2014) in minority ethnic groups seem to extend to the level of symptom networks. Here, they appear to be expressed as an increased co-occurrence of hallucinations and persecutory ideations in men from a minority ethnic background compared to men from the majority White or Mixed ethnic background. This finding agrees with the idea that, under the influence of risk factors, hallucinations and delusions can become connected, which has been linked to worse prognosis and symptom persistence (Binbay et al., 2012; Smeets et al., 2012; Smeets, Lataster, Viechtbauer, Delespaul, & G.R.O.U.P., 2014; van Os & Reininghaus, 2016). Taken together with the present results, this may reinforce evidence that demonstrates that people from a minority ethnic background, particularly men, are at increased risk for poor mental health outcomes (Morgan et al., 2017; Singh et al., 2015). With the present data, however, it cannot be excluded that ethnicity acts as a proxy measure for factors not covered by our analysis, such as specific forms of deprivation. Delineating how mental health outcomes in men from a minority ethnic background are determined is an outstanding task for future research and may help to design more effective interventions. Identifying potential commonalities underlying minority ethnic group status and domestic violence, both of which were associated with increased co-occurrence of psychotic experiences in men and women, respectively, may prove insightful in this context.

Except for domestic violence, which was a relevant moderating factor in women and men, different risk factors explained heterogeneity in symptom networks of women and men, suggesting a likely complex interplay between sex and risk factors in impacting psychosis risk. Childhood sexual and physical abuse, for instance, were sources of heterogeneity in symptom networks of women, but not men. This finding adds to previous research suggesting particularly detrimental effects of sexual and physical abuse on mental health of girls and women (Adams, Mrug, & Knight, 2018; Thompson, Kingree, & Desai, 2004). One reason for the distinct role of adversities may lie in the sex-specific effects they have on the nervous system, against the backdrop of sex differences in maturation, structure and functioning thereof (DeSantis et al., 2011; Dow-Edwards, 2020; Hodes & Epperson, 2019; Popovic et al., 2020). Moreover, characteristics of some risk factors have been shown to differ by sex: men are more likely to engage in more escalating and chronic patterns of cannabis use than women, for example (Hawes, Trucco, Duperrouzel, Coxe, & Gonzalez, 2019; Wagner & Anthony, 2007). Girls, on the other hand, are more likely than boys to experience severe forms of sexual abuse within close victim-perpetrator relationships (Gold, Elhai, Lucenko, Swingle, & Hughes, 1998; Kendall-Tackett, Williams, & Finkelhor, 1993). Such variations may contribute to differing patterns of relationships between risk and symptom expression in women and men.

Overall, our results corroborate a growing realization that research should appraise that mechanisms contributing to psychosis expression may, at least in parts, differ by sex (Hodes & Epperson, 2019; Riecher-Rössler et al., 2018; Rosen et al., 2019; Stainton et al., 2021). As clinical research works towards early identification and individually tailored preventive interventions, the complex interplay between sex and environmental factors in impacting psychosis risk needs to be better understood to optimize these efforts (Hartung & Lefler, 2019; Riecher-Rössler et al., 2018; Rosen et al., 2019; Stainton et al., 2021). This includes disaggregating results by sex and gender in psychosis research more consistently (Hartung & Lefler, 2019), for example by documenting differences and similarities in symptom networks of women and men.

## Limitations

Results from the present study should be interpreted given several limitations. First, posterior predictive checks used for comparing the overall network connectivity tend to be conservative (Williams et al., 2020), which may have resulted in low sensitivity in post-hoc comparisons. This factor, and small sample sizes in some subgroups, may explain why we found no evidence that exposure to risk factors was associated with more densely connected symptom networks compared to non-exposure, contrary to our hypothesis. Effects of risk factors on symptom networks seem to be more specific, impacting single relations between symptoms rather than connectivity between all symptoms. Second, model-based recursive partitioning identifies those variables that reduce heterogeneity in symptom networks the most. Thus, age of the respondent, alcohol use, separation experiences, bullying and socioeconomic deprivation may explain heterogeneity in symptom networks, but not to the same extent as the other risk factors tested. Related, differential relevance of risk factors, for example within ethnic groups, may have remained undetected due to small sample sizes in some subgroups. For a better understanding of the mechanisms relevant in different minority groups, targeted investigations in these populations with larger sample sizes are needed. Third, we did not incorporate complex design features of the APMS, such as weights to take non-response into account, due to the lack of established methods to do so for network models (Lin et al., 2019). Related, recursive partitioning currently only allows for complete case analyses. Even though the percentage of excluded participants was small, they differed from included participants in some important aspects, including hallucinatory and depressive symptoms, which may have biased our results. While therefore not fully representative of the English population, our results are based on a large national household survey, with suitability for a data-intensive method, such as network-based recursive partitioning, unlike for smaller samples which would not offer the same opportunity. Fourth, the retrospective assessment of risk factors via self-report may be prone to memory biases and so directions of effect may be contested. Fifth, data used in the present analyses were gathered in a large epidemiological study; therefore, instruments and tools used were designed such that they were simple to understand and appropriate given their use in over 7,000 people. This setting necessarily leads to less refined assessments of symptomatology and risk. Sixth, the analyses were based on cross-sectional data, meaning that the directions of interactions among the symptoms remain unknown. Longitudinal studies are therefore an important next step for this line of research, and extension of recursive partitioning methods to personalized network structures (e.g., derived from experience sampling methods) may allow for insights into how risk factors moderate dynamic associations between symptoms in individuals. Lastly, some researchers have expressed concerns about stability and replicability of network models (e.g., estimates of edges (Forbes, Wright, Markon, & Krueger, 2017); for a summary of the debate, see McNally, 2021). While our robustness analysis suggests that the networks and edge estimates are generally stable, especially weaker links in the networks of small subgroups should be interpreted with care. Given that recursive partitioning and network methodology are data-driven, replication of present findings in other samples is needed to establish generalizability (Fried et al., 2018).

## Conclusion

Symptom networks of psychosis expression in the general population are highly heterogeneous. Sex was the primary source of heterogeneity, and different risk factors explained further variability in symptom networks of women and men, potentially reflecting distinct sex-specific mechanisms contributing to psychosis risk. Among women, an affective loading within psychosis, particularly following early interpersonal trauma, may have distinct importance. Among men, the symptom network of those from a minority ethnic background showed a particularly strong connection between hallucinatory experiences and persecutory ideation, which may reflect poorer outcomes including symptom resolution in this group. A better understanding and consideration of these sex differences provides an important opportunity to deliver high quality prevention and patient-centered care in psychosis.

## Data Availability

Data of the 2007 APMS (National Centre for Social Research, University of Leicester, 2017) used in the analyses are available from the UK Data Service (https://ukdataservice.ac.uk/).

## Acknowledgements

None.

## Required Statements

### Financial Support

JK has received funding from the German Research Foundation (DFG; grant agreement n° KA 4413/1-1). MR has received funding from the Köln Fortune program (grant agreement n° 304/2020).

### Conflict of Interest

The authors declare no conflict of interests with relation to the work reported in this manuscript.

### Ethical Standards

The authors assert that all procedures contributing to this work comply with the ethical standards of the relevant national and institutional committees on human experimentation and with the Helsinki Declaration of 1975, as revised in 2008.

## Notes

### Competing Interest Statement

The authors have declared no competing interest.

### Funding Statement

JK has received funding from the German Research Foundation (DFG; grant agreement number KA 4413/1-1). MR has received funding from the Koeln Fortune program (grant agreement number 304/2020).

### Author Declarations

Ethical approval for APMS 2007 was obtained from the Royal Free Hospital and Medical School Research Ethics Committee.

### Summary of Updates

Methods updated to expand on estimation procedure and to add stability checks; results updated to include stability checks; discussion updated to provide a more nuanced view on the generalizability of network analyses; references added for clarity; supplemental files updated.

