## Supplementary Materials for "Disentangling heterogeneity of psychosis expression in the general population: sex-specific moderation effects of environmental risk factors on symptom networks"

**Supplementary method 1.** Assessment of psychopathology.

**Supplementary method 2.** Assessment of demographic and environmental risk factors.

**Supplementary method 3.** Identification of network subgroups via recursive partitioning.

**Supplementary table 1.** Comparison of participants included in and excluded from the analysis due to missing values.

**Supplementary table 2.** Results from case-drop subset bootstrapping.

**Supplementary figure 1**. Results from recursive partitioning disaggregated by sex, depicted as decision trees of partial correlation networks.

**Supplementary figure 2.** Accuracy of the edge-weights in the network estimated in the full sample.

**Supplementary figure 3.** Accuracy of the edge-weights in the network estimated in women.

**Supplementary figure 4.** Accuracy of the edge-weights in the network estimated in men.

**Supplementary figure 5.** Accuracy of the edge-weights in the network estimated in women who reported sexual abuse in childhood.

**Supplementary figure 6.** Accuracy of the edge-weights in the network estimated in women who did not report sexual abuse in childhood.

**Supplementary figure 7.** Accuracy of the edge-weights in the network estimated in women who did not report sexual abuse in childhood, but physical abuse.

**Supplementary figure 8.** Accuracy of the edge-weights in the network estimated in women who reported neither sexual abuse nor physical abuse in childhood.

**Supplementary figure 9.** Accuracy of the edge-weights in the network estimated in women who reported neither sexual abuse nor physical abuse in childhood, but domestic violence.

**Supplementary figure 10.** Accuracy of the edge-weights in the network estimated in women who reported neither sexual abuse in childhood, physical abuse in childhood, nor domestic violence.

**Supplementary figure 11.** Accuracy of the edge-weights in the network estimated in men who reported domestic violence.

**Supplementary figure 12.** Accuracy of the edge-weights in the network estimated in men who did not report domestic violence.

**Supplementary figure 13.** Accuracy of the edge-weights in the network estimated in men who did not report domestic violence, but cannabis use in the past year.

**Supplementary figure 14.** Accuracy of the edge-weights in the network estimated in men who reported neither domestic violence nor cannabis use in the past year.

**Supplementary figure 15.** Accuracy of the edge-weights in the network estimated in men who reported neither domestic violence nor cannabis use and reported having a White or ‘Other’ ethnic background.

**Supplementary figure 16.** Accuracy of the edge-weights in the network estimated in men who reported neither domestic violence nor cannabis use and reported having a Black or South Asian ethnic background.

**Supplementary methods**

**Supplementary method 1.** Assessment of psychopathology.

We obtained affective symptoms of worry, sleep disturbance, generalized anxiety and depression from the Revised Version of the Clinical Interview Schedule (CIS-R; Lewis, Pelosi, Araya, & Dunn, 1992). We coded these symptoms as present if they had been reported in the past month and persisted for at least two weeks (Moffa et al., 2017). Persecutory ideation and hallucinatory experiences were obtained from the Psychosis Screening Questionnaire (Bebbington & Nayani, 1995). Persecutory ideation was defined as present given a positive response to the question: ‘Over the past year, have you felt that people were deliberately acting to harm you/your interests?’. Similarly, hallucinatory experiences were defined as a positive response to the question: ‘In the past year, have you ever heard voices saying quite a few words or sentences when there was no-one around that might explain it?’

**Supplementary method 2.** Assessment of demographic and environmental risk factors.

**Early-life adversities*.*** Early-life adversities comprised physical abuse and sexual abuse before the age of 16, as well as separation experiences from parents (institutional care and local authority care until the age of 16).

Specifically, physical abuse was defined as a positive response to the question: ‘Before the age of 16, were you ever severely beaten by a parent, step-parent or carer?’. Sexual abuse was defined via a positive endorsement of any of the following questions: ‘Before the age of 16, did anyone talk to you in a sexual way that made you feel uncomfortable?’, ‘Before the age of 16, did anyone touch you, or get you to touch them, in a sexual way without your consent?’, and ‘Before the age of 16, did anyone have sexual intercourse with you without your consent?’.

Two items taken from the ‘parenting’ section of the APMS (National Centre for Social Research, University of Leicester, 2017) were selected to define separation from parents, requiring “yes” responses to at least one of the following questions: Institutional care: ‘Up to the age of 16 did you spend any time in any kind of institution such as a children’s home, borstal, or young offenders unit?’; Local Authority care: ‘Were you ever taken into Local Authority care (i.e., into a children’s home or foster care) as a child up until the age of 16?’

**Later-life adversities.** Later-life adversities comprised bullying, domestic violence, cannabis use in the past year, alcohol use, ethnic background, and deprivation.

Specifically, respondents had to select ‘bullying’ and/or ‘violence in the home’ from a list of options on a card following the question, ‘Now looking at this card, could you tell me if you have ever experienced any of these problems or events, at any time in your life?’. We coded responses as “yes” if the respective risk factor was reported and “no” if not.

Cannabis use in the past year was defined via a positive response to the question ‘Have you used cannabis in the last year?’.

Alcohol use was assessed with the Alcohol Use Disorders Identification Test (AUDIT; Saunders, Aasland, Babor, de La Fuente, & Grant, 1993). The range of possible scores is from 0 to 40 where 0 indicates an abstainer who has never had any problems from alcohol. A score of 1 to 7 suggests low-risk consumption according to World Health Organization (WHO) guidelines. Scores from 8 to 14 suggest hazardous or harmful alcohol consumption and a score of 15 or more indicates the likelihood of alcohol dependence (moderate-severe alcohol use disorder).

Respondents were also asked to indicate their ethnicity, and our analyses included four categories: White, Black, South Asian and Mixed/Other.

Deprivation in the APMS 2007 was assessed via index of multiple deprivation (IMD) 2004 data. This is a derived measure of social and economic deprivation based on seven domains or neighborhood variables: income; employment; health and disability; education, skills and training; barriers to housing and services; living environment; and crime. Data for the IMD 2004 was collected between 1997 and 2003 and the APMS 2007 (National Centre for Social Research, University of Leicester, 2017) reported 5 quintiles of deprivation, where 1 represents the least deprived and 5 represents most deprived.

**Supplementary method 3.** Identification of network subgroups via recursive partitioning.

We used a model-based recursive partitioning approach to identify meaningful subgroups given the included environmental and demographic factors, as implemented in the R package ‘networktree’, version 1.0.1 (Jones, Mair, Simon, & Zeileis, 2020). The ‘networktree’ approach determines sample splits based on significant invariance in the correlation matrix of the network variables under consideration, yielding non-overlapping partitions of the sample with maximally heterogeneous symptom networks (Jones et al., 2020). Specifically, a log-likelihood-based score function is used to assess how much each participant deviates from parameters (i.e., pairwise correlation-coefficients in the correlation matrix that are the basis for network models) estimated in the full sample. In a well-fitting symptom network model not subject to significant heterogeneity, individual deviations should be close to 0 and randomly fluctuate around it. Based on structural change tests, p-values are generated for each moderating variable, assessing the extent to which a given moderating variable can capture deviations in parameters, i.e., heterogeneity in symptom networks. The moderating variable with the lowest p-value serves as a so-called splitting variable, given that this p-value is below α, which is Bonferroni-corrected for the number of moderating variables tested (Jones et al., 2020). Thus, the algorithm prioritizes the “biggest” splits first. The procedure of testing for invariance in network structures is repeated recursively within the newly identified subgroups until no significant invariance in network structures can be detected. To ensure stability and interpretability of the results, we set the minimum number of observations within any final group of the decision tree to 1% of the total sample size, i.e., 73.

**Supplementary tables**

**Supplementary table 1.** Comparison of participants included in and excluded from the analysis due to missing values. We used permutation-tests as implemented in the *R* package ‘coin’ (Hothorn, Hornik, van de Wiel, & Zeileis, 2008), specifically the Pearson χ^2-^ test for count data, and the Wilcoxon-test for interval data.

| Variable  Yes (%)/Median (IQR) | Included  (*n* = 7,242) | Excluded  (*n* = 161) | Test statistic, *p*-value |
| --- | --- | --- | --- |
| Network Variables |  |  |  |
| worry | 36.0 | 33.8 | χ^2^ = 0.32, *p* = .609 |
| sleep problems | 34.6 | 41.6 | χ^2^ = 3.37, *p* = .082 |
| anxiety | 17.3 | 18.6 | χ^2^ = 0.21, *p* = .676 |
| depression | 22.9 | 35.0 | χ^2^ = 12.8, *p* < .001 |
| persecutory ideation | 7.7 | 7.7 | χ^2^ = 0.001, *p* = 1 |
| hallucinatory experiences | 0.80 | 0.50 | χ^2^ = 22.5, *p* < .001 |
| Potential Moderators | | | |
| sex (% female) | 56.8 | 56.5 | χ^2^ = 0.006, *p* = 1 |
| age (years) | 50 (30) | 58 (40) | *Z* = 4.53, *p* < .001 |
| ethnic background | White: 92.7, Black: 2.6, South Asian: 2.6, Mixed/Other: 2.1 | White: 85.6, Black: 1.8, South Asian: 7.2, Mixed/Other: 5.4 | χ^2^ = 14.8, *p* = .006 |
| deprivation | 3 (2) | 3 (3) | *Z* = 2.76, *p =* .005 |
| bullying | 18.9 | 19.1 | χ^2^ = 0.003, *p* = 1 |
| separation from parents | 3.4 | 4.2 | χ^2^ = 0.21, *p* = .803 |
| domestic violence | 9.5 | 8.7 | χ^2^ = 0.08, *p* = .877 |
| physical abuse | 4.8 | 8.5 | χ^2^ = 2.09, *p* = .151 |
| sexual abuse | 13.5 | 10.8 | χ^2^ = 0.41, *p* = .595 |
| cannabis use in past year | 5.7 | 2.6 | χ^2^ = 2.00, *p* = .212 |
| alcohol consumption  (AUDIT score) | 4 (5) | 2 (5) | *Z* = -4.60, *p* < .001 |

**Supplementary table 2.** Results from case-drop subset bootstrapping (Costenbader & Valente, 2003; Epskamp, Borsboom, & Fried, 2018). For subgroup networks, see figure 2 in main manuscript.

| Subgroup | CS coefficient |
| --- | --- |
| Full sample | 0.75 |
| *First split (figure 2a)* |  |
| women | 0.75 |
| men | 0.75 |
| *Second split (figure 2b)* |  |
| sexual abuse: yes | 0.75 |
| sexual abuse: no | 0.75 |
| *Third split (figure 2c)* |  |
| physical abuse: yes | 0.28 |
| physical abuse: no | 0.75 |
| *Fourth split (figure 2d)* |  |
| domestic violence: yes | 0.52 |
| domestic violence: no | 0.75 |
| *Fifth split (figure 2e)* |  |
| domestic violence: yes | 0.44 |
| domestic violence: no | 0.75 |
| *Fifth split (figure 2f)* |  |
| cannabis: yes | 0.44 |
| cannabis: no | 0.75 |
| *Sixth split (figure 2g)* |  |
| ethnicity: Black or South Asian | 0.52 |
| ethnicity: White or Mixed/Other | 0.75 |

*Abbreviations*: CS coefficient = correlation stability coefficient.

*Note*: Figure 2 can be found in the main manuscript.

**Supplementary figures**

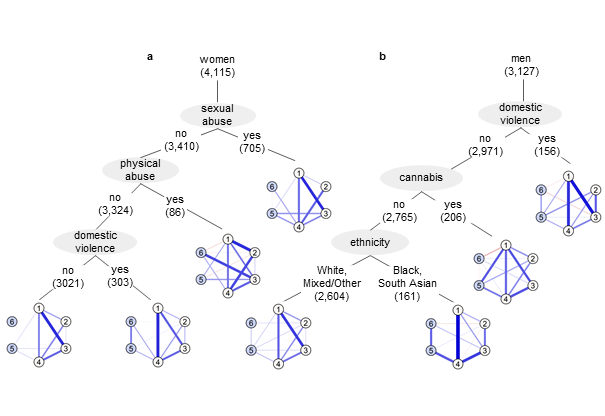

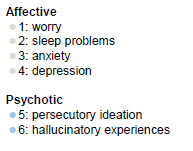

**Supplementary figure 1.** Results from recursive partitioning disaggregated by sex, depicted as decision trees of partial correlation networks. Numbers behind splitting factors give the sample size retained after the corresponding sample split. Symptom groups are differentiated by color. The thicker and less transparent the edge, the stronger the partial correlation between two symptoms. Blue (red) edges indicate positive (negative) relationships. To ensure visual comparability, edge weights were scaled identically across all networks. Only edge weights larger than 0.01 are depicted.

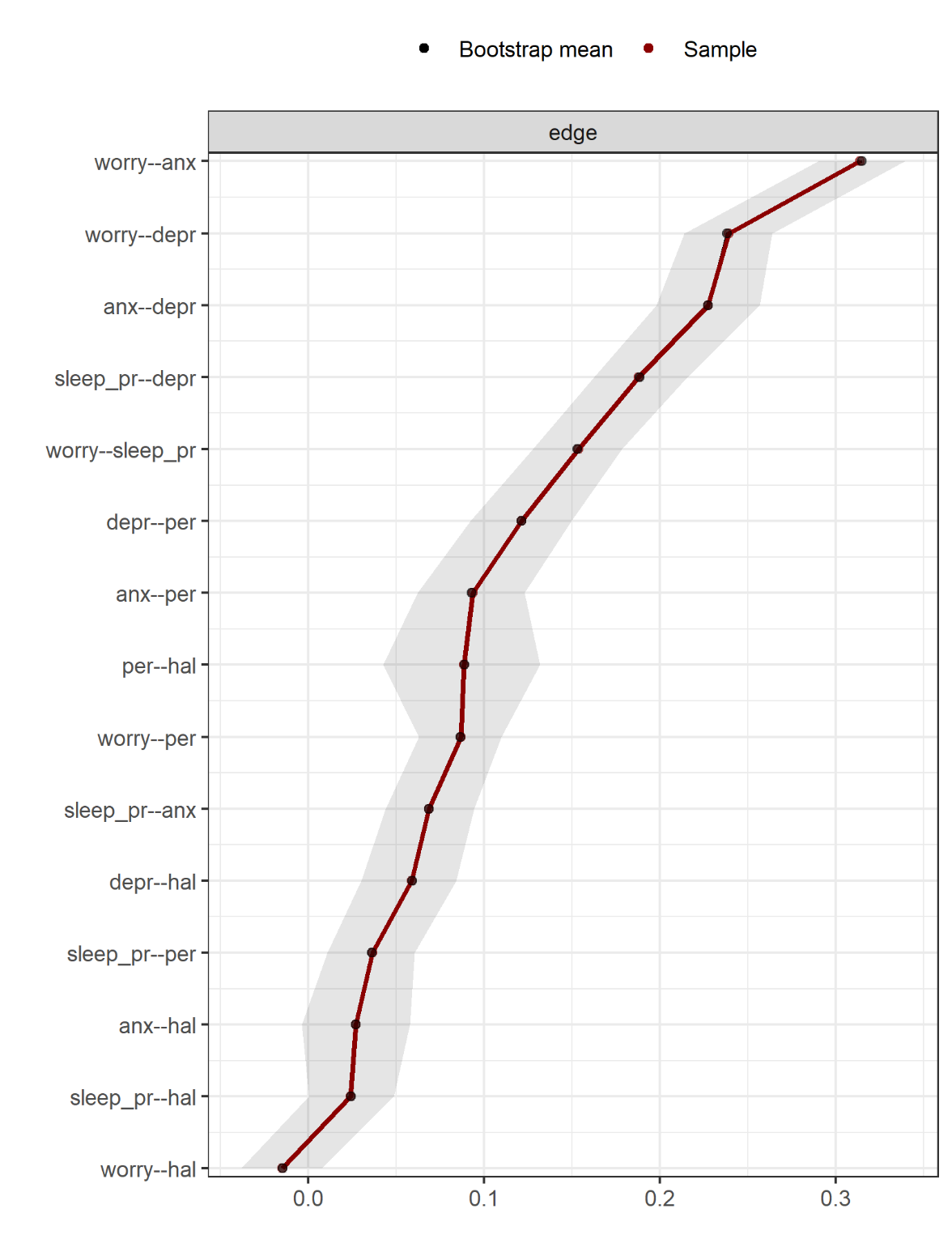

**Supplementary figure 2.** Accuracy of the edge-weights in the network estimated in the full sample. The grey horizonal area within the plot represents the 95% quantile range of the parameter values across 5000 bootstraps. The red dots indicate the sample values for the analyzed data, while the black dots indicate the bootstrap mean values. The sample values lie within the bootstrapped confidence intervals and the bootstrap mean values are very well-aligned with the sample values, thus indicating accurate estimations.

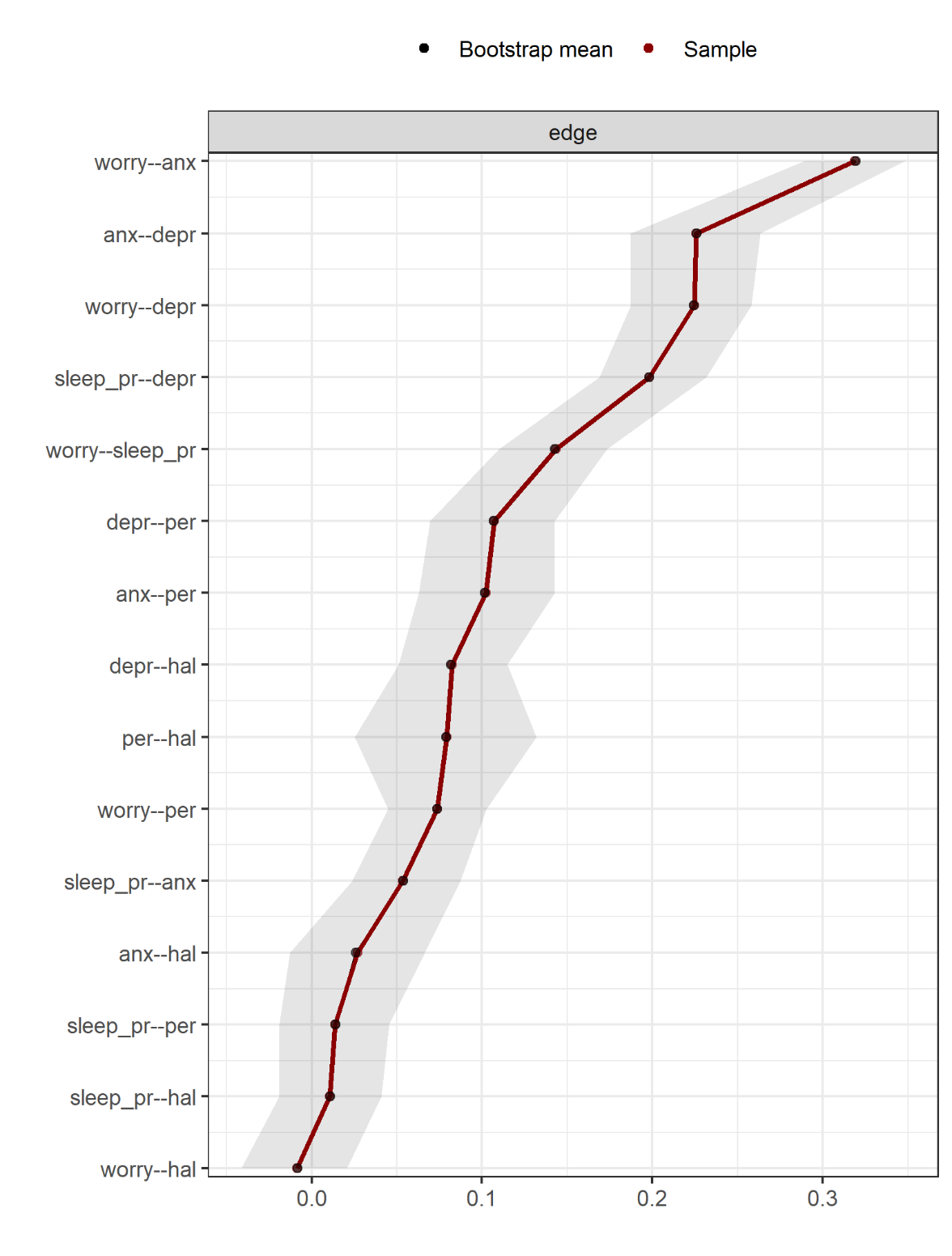

**Supplementary figure 3.** Accuracy of the edge-weights in the network estimated in women. The grey horizonal area within the plot represents the 95% quantile range of the parameter values across 5000 bootstraps. The red dots indicate the sample values for the analyzed data, while the black dots indicate the bootstrap mean values. The sample values lie within the bootstrapped confidence intervals and the bootstrap mean values are very well-aligned with the sample values, thus indicating accurate estimations.

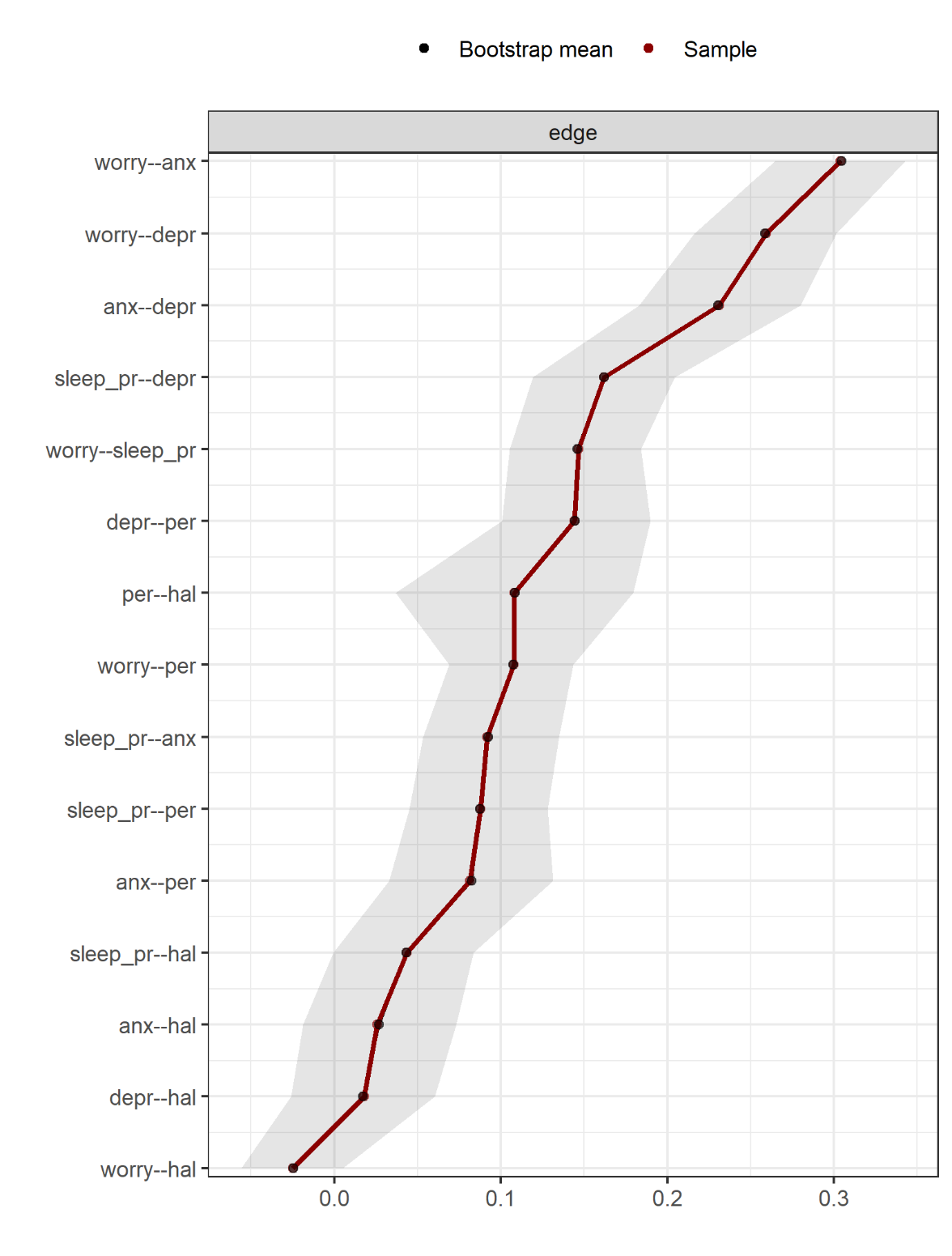

**Supplementary figure 4.** Accuracy of the edge-weights in the network estimated in men. The grey horizonal area within the plot represents the 95% quantile range of the parameter values across 5000 bootstraps. The red dots indicate the sample values for the analyzed data, while the black dots indicate the bootstrap mean values. The sample values lie within the bootstrapped confidence intervals and the bootstrap mean values are very well-aligned with the sample values, thus indicating accurate estimations.

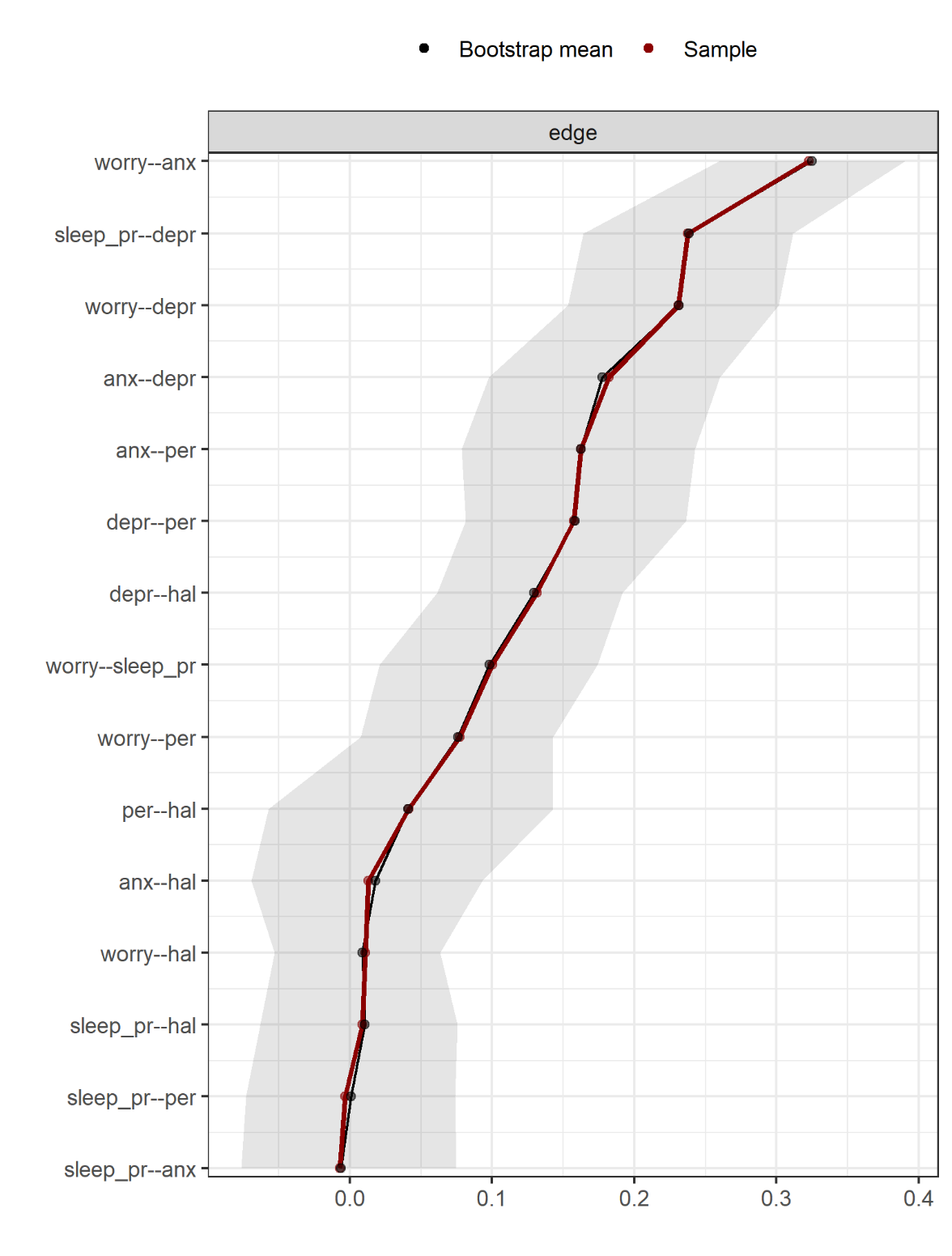

**Supplementary figure 5.** Accuracy of the edge-weights in the network estimated in women who reported sexual abuse in childhood. The grey horizonal area within the plot represents the 95% quantile range of the parameter values across 5000 bootstraps. The red dots indicate the sample values for the analyzed data, while the black dots indicate the bootstrap mean values. The sample values lie within the bootstrapped confidence intervals and the bootstrap mean values are very well-aligned with the sample values, thus indicating accurate estimations.

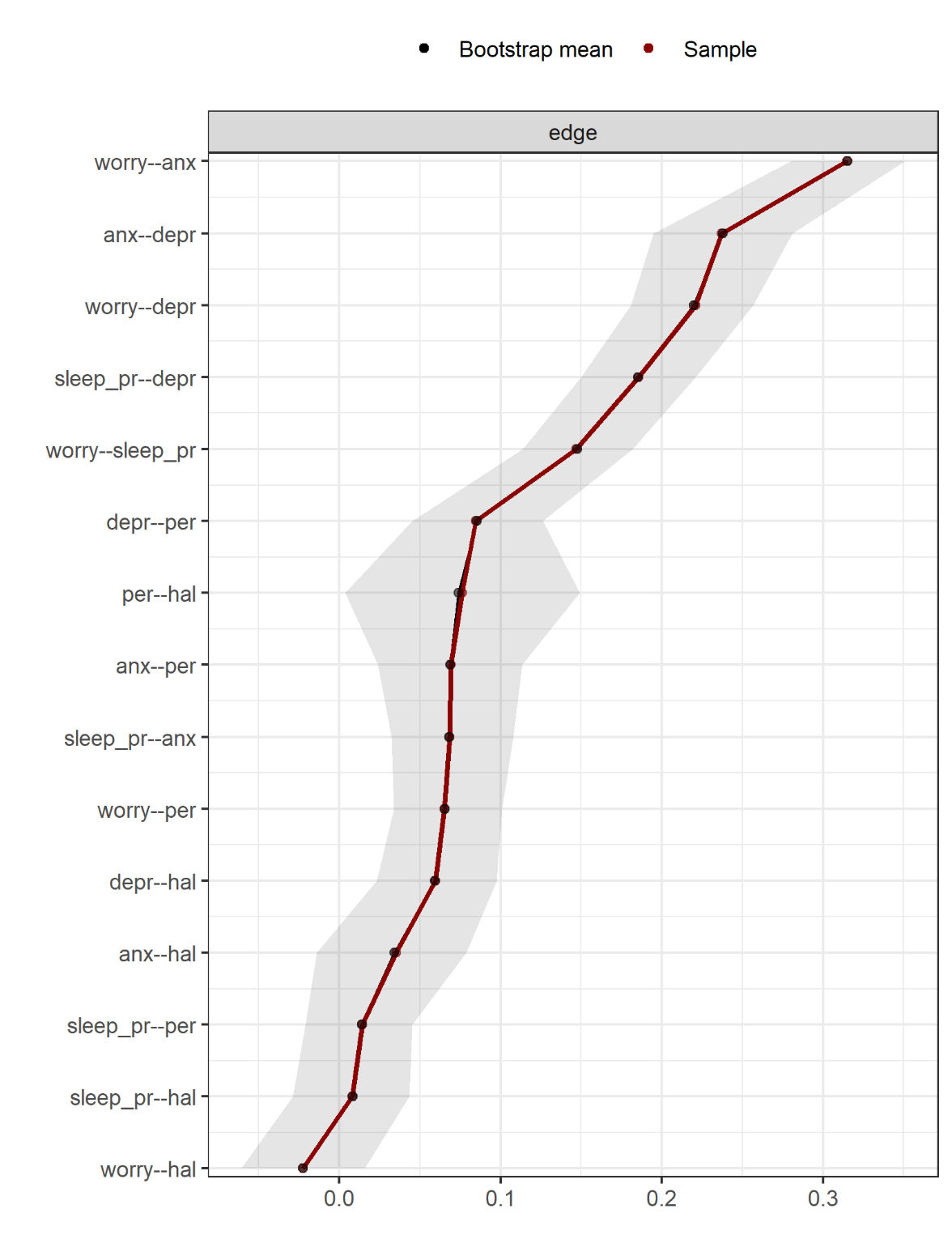

**Supplementary figure 6.** Accuracy of the edge-weights in the network estimated in women who did not report sexual abuse in childhood. The grey horizonal area within the plot represents the 95% quantile range of the parameter values across 5000 bootstraps. The red dots indicate the sample values for the analyzed data, while the black dots indicate the bootstrap mean values. The sample values lie within the bootstrapped confidence intervals and the bootstrap mean values are very well-aligned with the sample values, thus indicating accurate estimations.

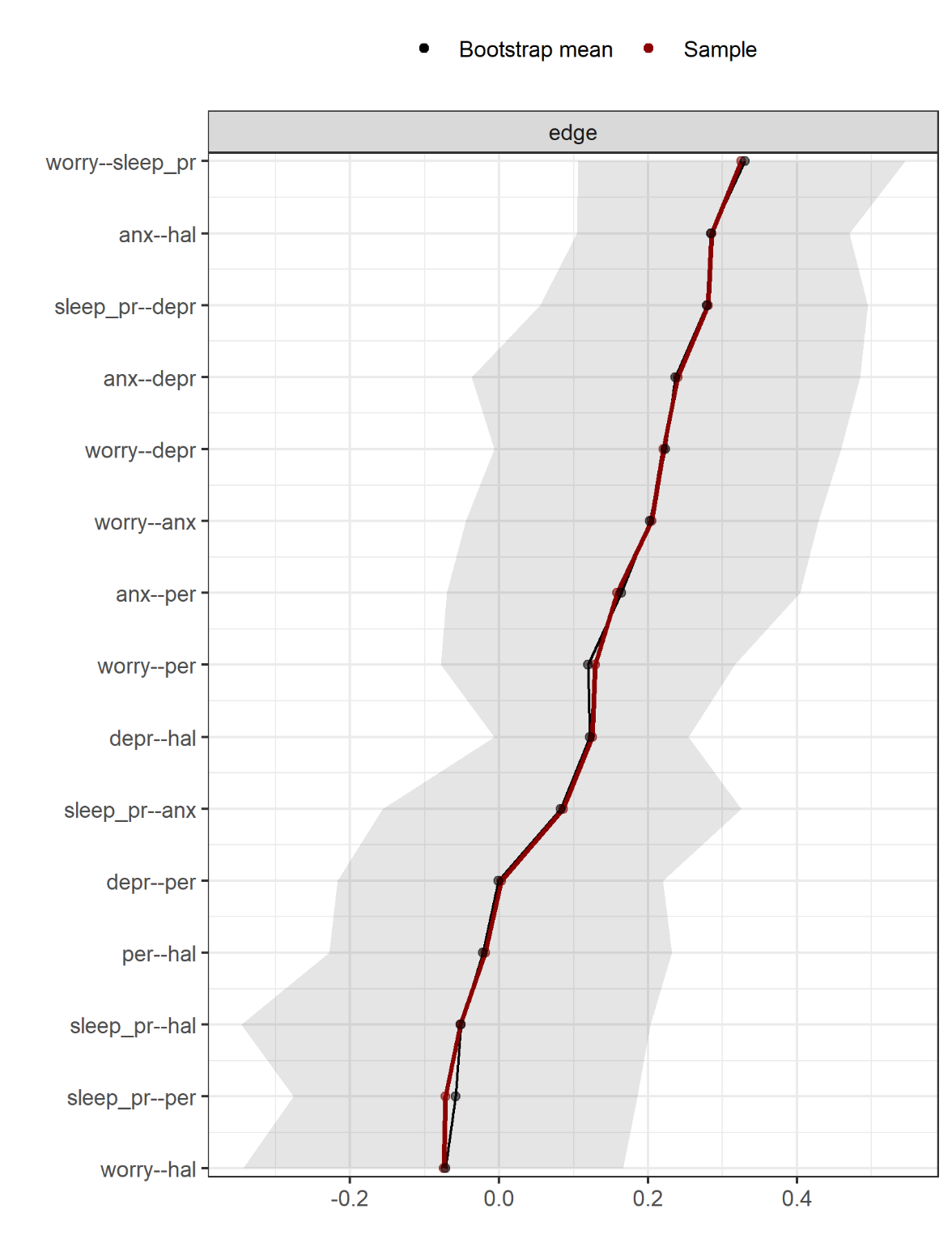

**Supplementary figure 7.** Accuracy of the edge-weights in the network estimated in women who did not report sexual abuse in childhood, but physical abuse. The grey horizonal area within the plot represents the 95% quantile range of the parameter values across 5000 bootstraps. The red dots indicate the sample values for the analyzed data, while the black dots indicate the bootstrap mean values. The sample values lie within the bootstrapped confidence intervals and the bootstrap mean values are generally well-aligned with the sample values, thus indicating accurate estimations. Of note, the bootstrapped confidence intervals are relatively wide, thus some caution is recommended especially when interpreting the strength of weaker edges.

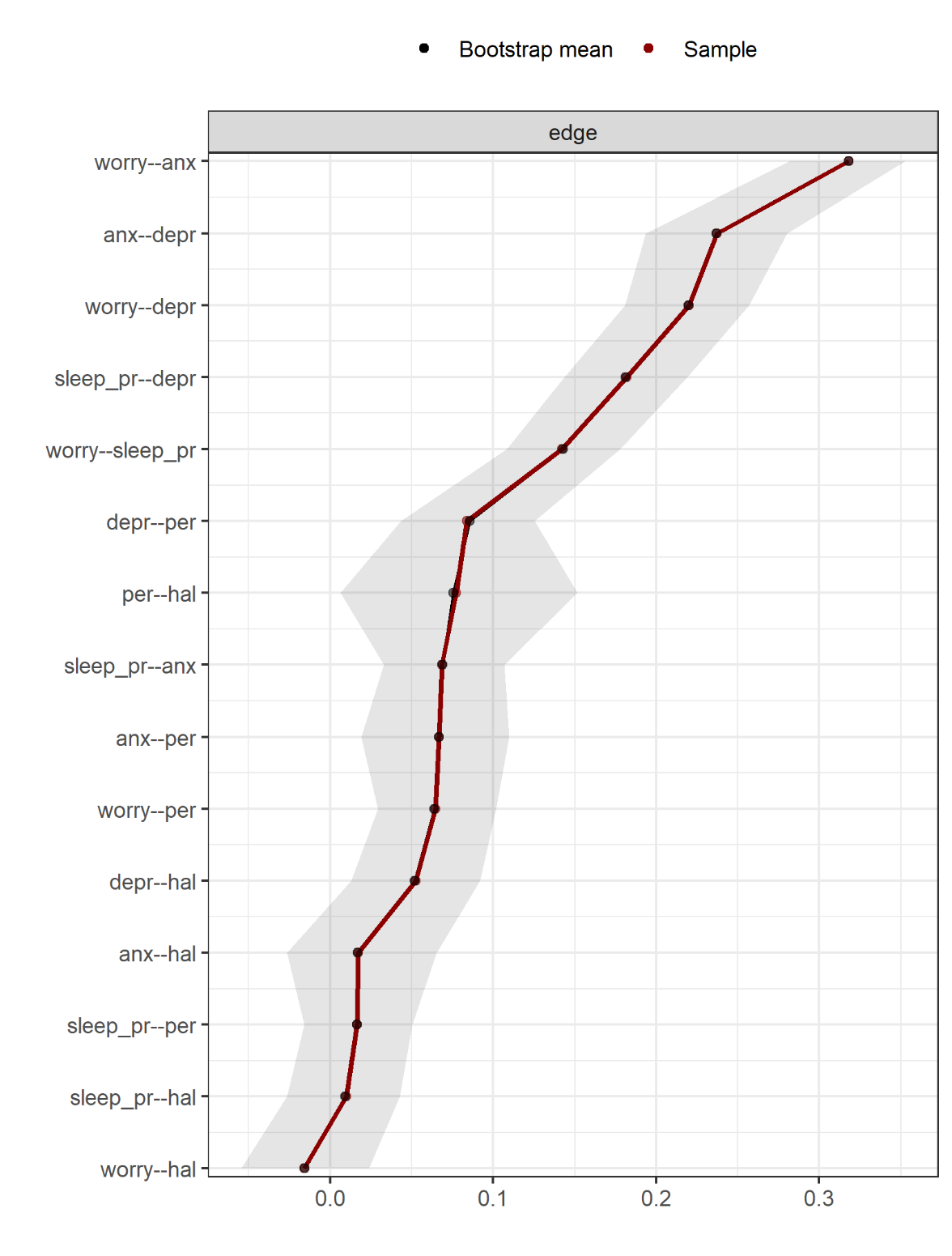

**Supplementary figure 8.** Accuracy of the edge-weights in the network estimated in women who reported neither sexual abuse nor physical abuse in childhood. The grey horizonal area within the plot represents the 95% quantile range of the parameter values across 5000 bootstraps. The red dots indicate the sample values for the analyzed data, while the black dots indicate the bootstrap mean values. The sample values lie within the bootstrapped confidence intervals and the bootstrap mean values are very well-aligned with the sample values, thus indicating accurate estimations.

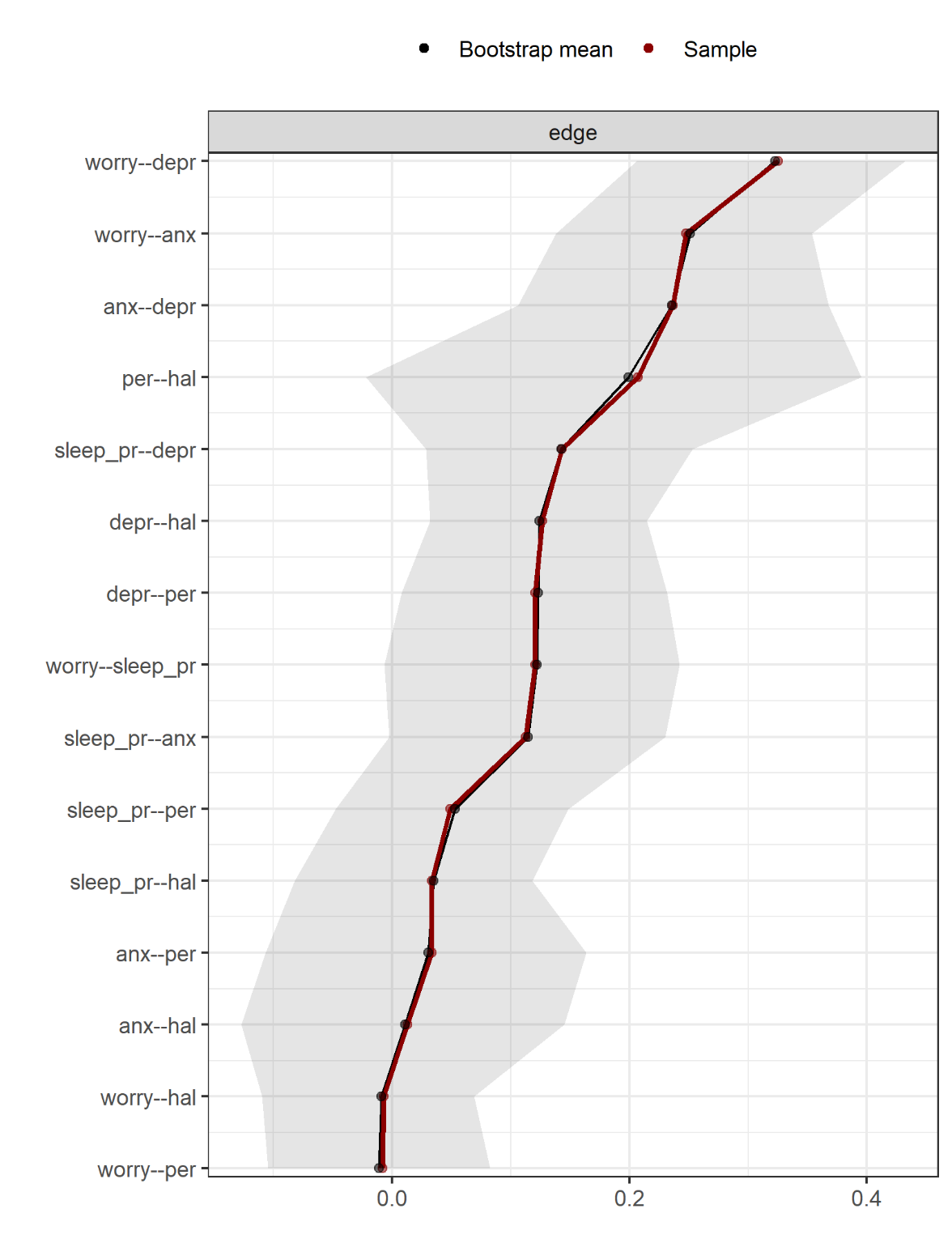

**Supplementary figure 9.** Accuracy of the edge-weights in the network estimated in women who reported neither sexual abuse nor physical abuse in childhood, but domestic violence. The grey horizonal area within the plot represents the 95% quantile range of the parameter values across 5000 bootstraps. The red dots indicate the sample values for the analyzed data, while the black dots indicate the bootstrap mean values. The sample values lie within the bootstrapped confidence intervals and the bootstrap mean values are generally well-aligned with the sample values, thus indicating accurate estimations. Of note, the bootstrapped confidence intervals are relatively wide, thus some caution is recommended especially when interpreting the strength of weaker edges.

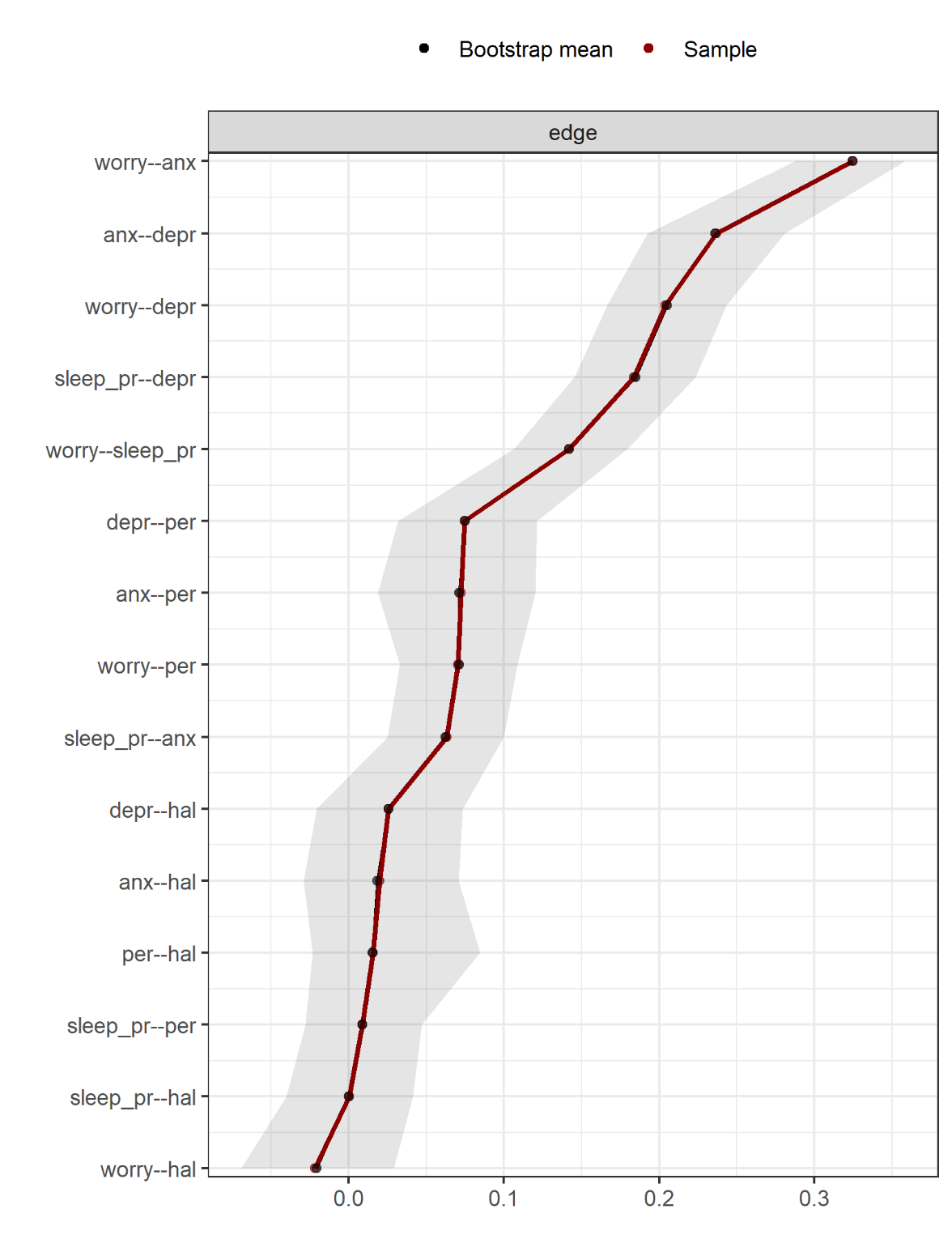

**Supplementary figure 10.** Accuracy of the edge-weights in the network estimated in women who reported neither sexual abuse in childhood, physical abuse in childhood, nor domestic violence. The grey horizonal area within the plot represents the 95% quantile range of the parameter values across 5000 bootstraps. The red dots indicate the sample values for the analyzed data, while the black dots indicate the bootstrap mean values. The sample values lie within the bootstrapped confidence intervals and the bootstrap mean values are very well-aligned with the sample values, thus indicating accurate estimations.

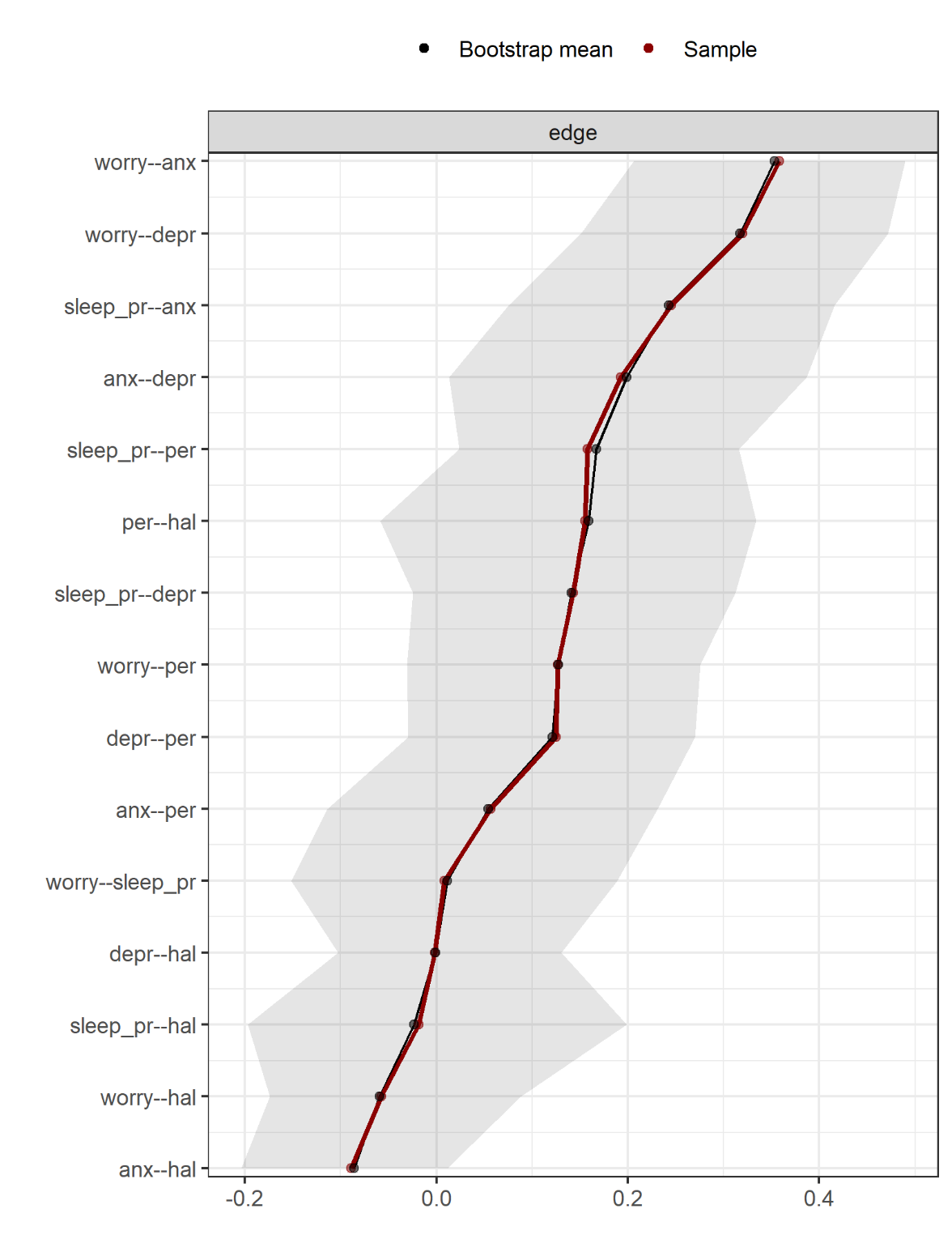

**Supplementary figure 11.** Accuracy of the edge-weights in the network estimated in men who reported domestic violence. The grey horizonal area within the plot represents the 95% quantile range of the parameter values across 5000 bootstraps. The red dots indicate the sample values for the analyzed data, while the black dots indicate the bootstrap mean values. The sample values lie within the bootstrapped confidence intervals and the bootstrap mean values are generally well-aligned with the sample values, thus indicating accurate estimations. Of note, the bootstrapped confidence intervals are relatively wide, thus some caution is recommended especially when interpreting the strength of weaker edges.

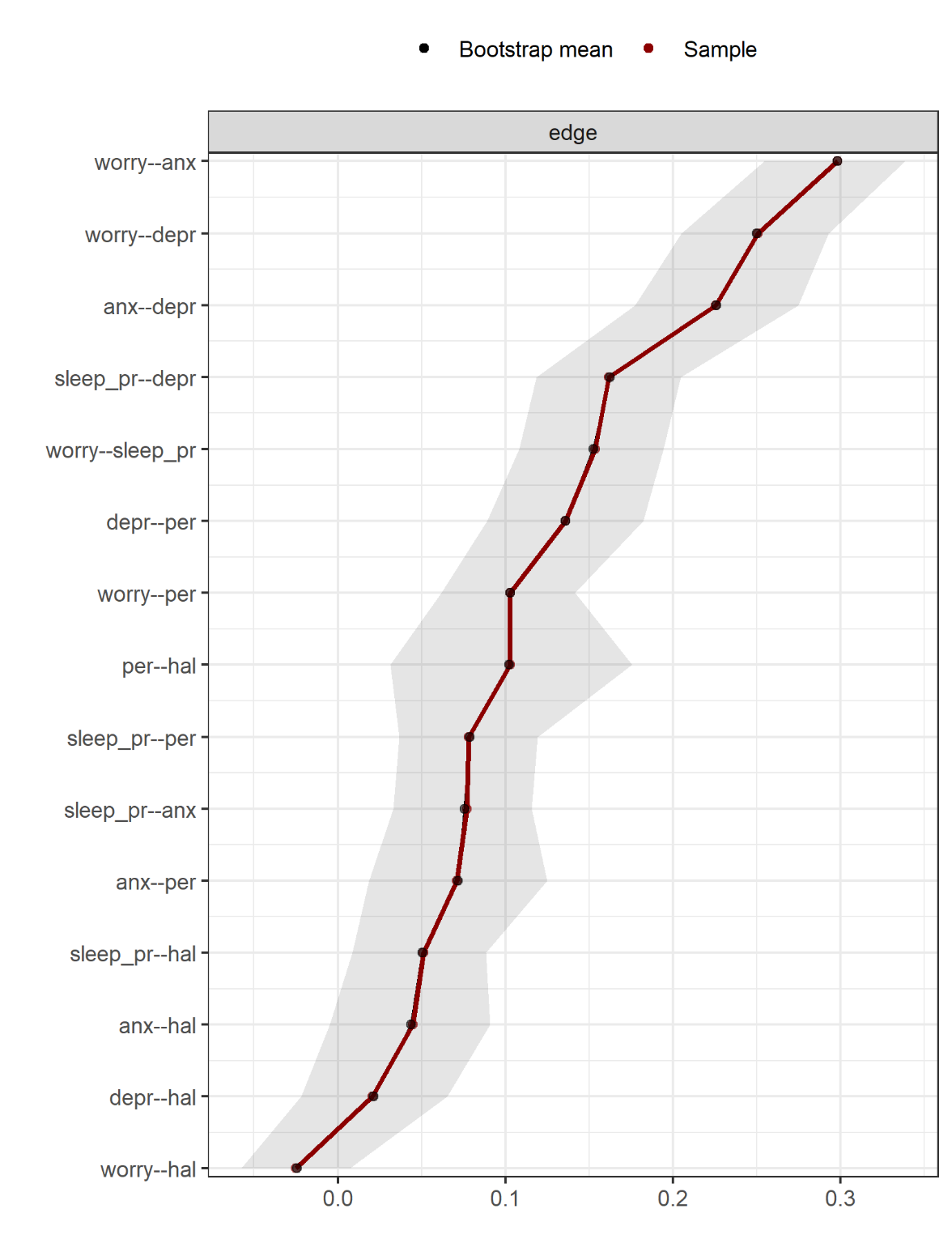

**Supplementary figure 12.** Accuracy of the edge-weights in the network estimated in men who did not report domestic violence. The grey horizonal area within the plot represents the 95% quantile range of the parameter values across 5000 bootstraps. The red dots indicate the sample values for the analyzed data, while the black dots indicate the bootstrap mean values. The sample values lie within the bootstrapped confidence intervals and the bootstrap mean values are very well-aligned with the sample values, thus indicating accurate estimations.

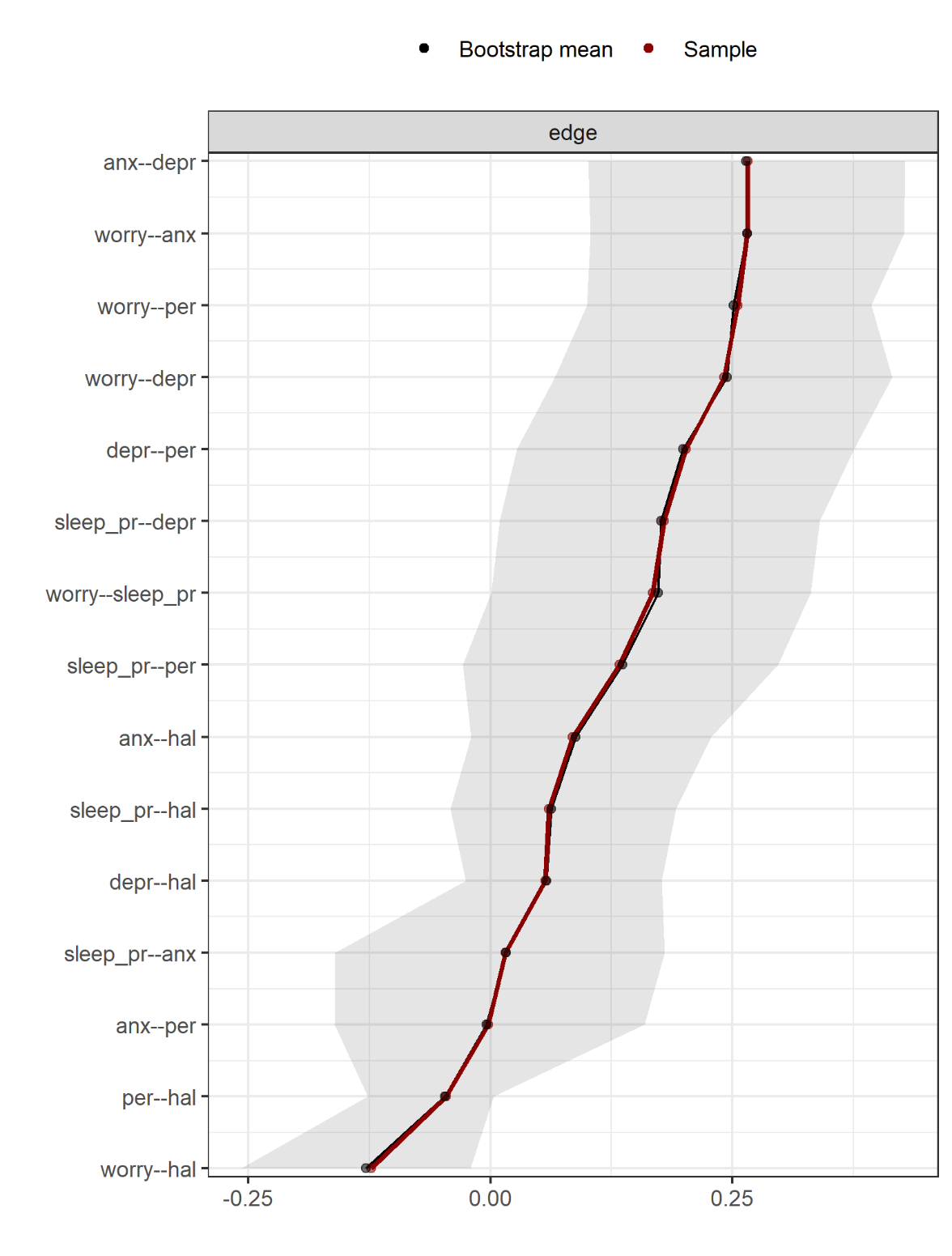

**Supplementary figure 13.** Accuracy of the edge-weights in the network estimated in men who did not report domestic violence, but cannabis use in the past year. The grey horizonal area within the plot represents the 95% quantile range of the parameter values across 5000 bootstraps. The red dots indicate the sample values for the analyzed data, while the black dots indicate the bootstrap mean values. The sample values lie within the bootstrapped confidence intervals and the bootstrap mean values are well-aligned with the sample values, thus indicating accurate estimations. Of note, the bootstrapped confidence intervals are relatively wide, thus some caution is recommended especially when interpreting the strength of weaker edges.

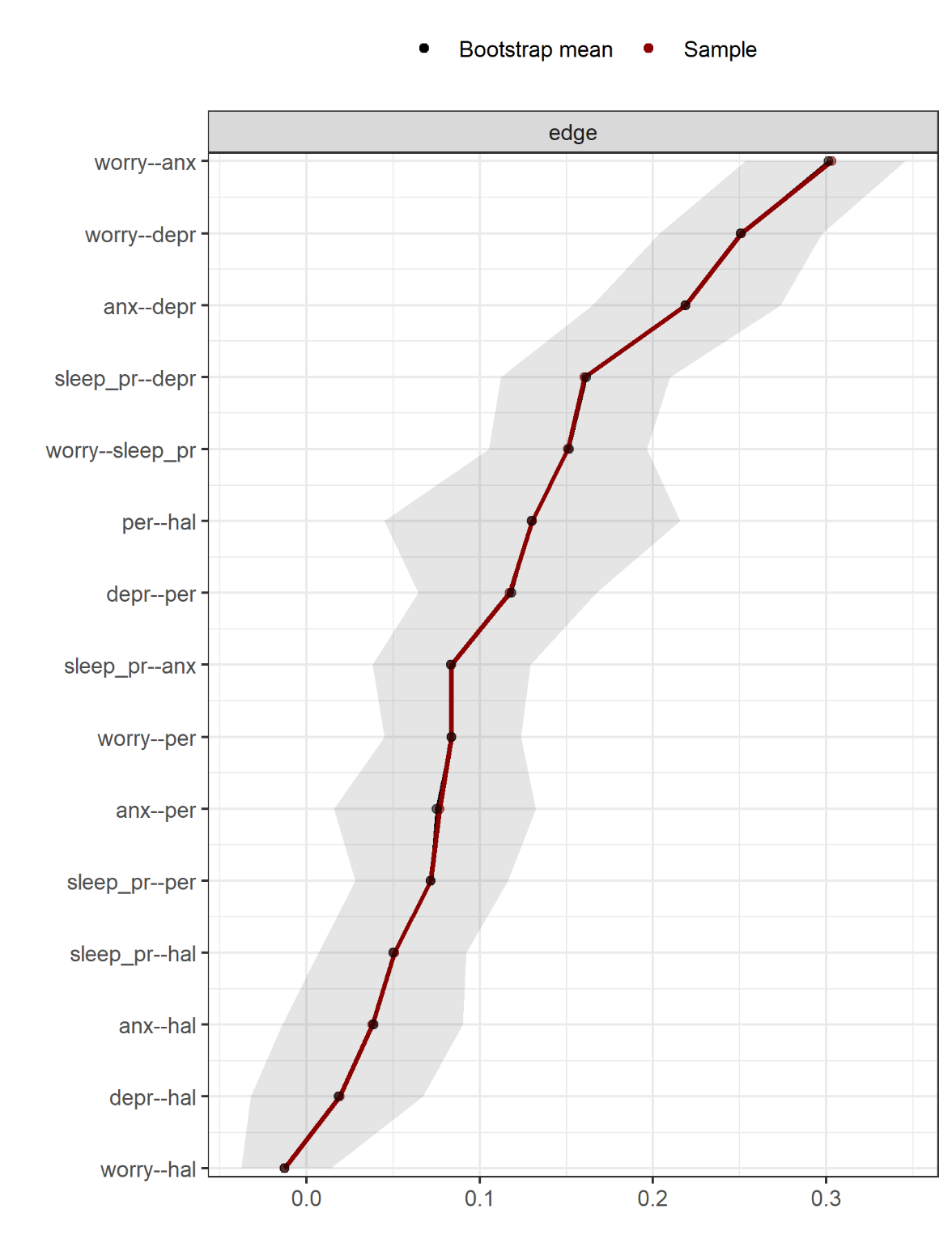

**Supplementary figure 14.** Accuracy of the edge-weights in the network estimated in men who reported neither domestic violence nor cannabis use in the past year. The grey horizonal area within the plot represents the 95% quantile range of the parameter values across 5000 bootstraps. The red dots indicate the sample values for the analyzed data, while the black dots indicate the bootstrap mean values. The sample values lie within the bootstrapped confidence intervals and the bootstrap mean values are very well-aligned with the sample values, thus indicating accurate estimations.

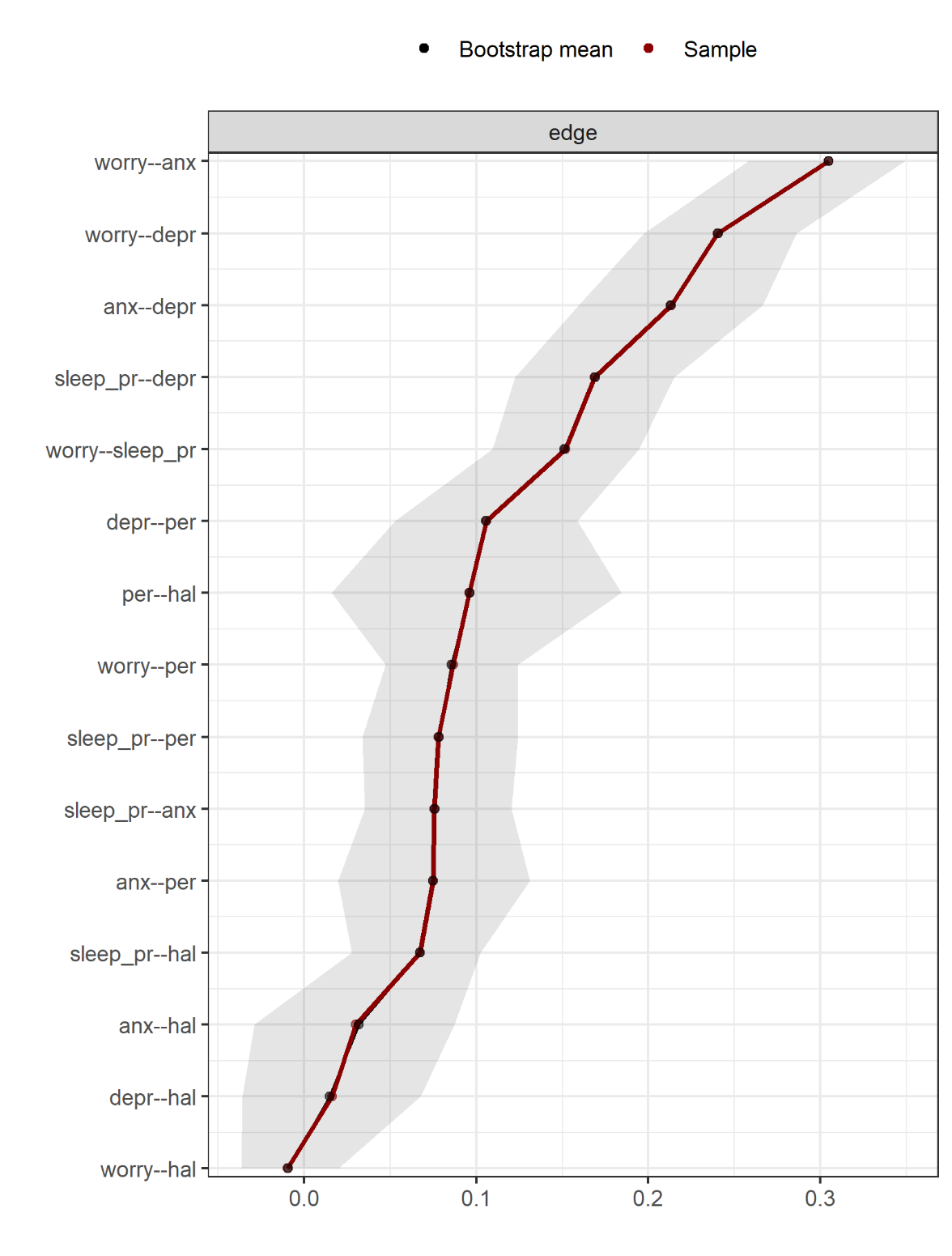

**Supplementary figure 15.** Accuracy of the edge-weights in the network estimated in men who reported neither domestic violence nor cannabis use and reported having a White or ‘Other’ ethnic background. The grey horizonal area within the plot represents the 95% quantile range of the parameter values across 5000 bootstraps. The red dots indicate the sample values for the analyzed data, while the black dots indicate the bootstrap mean values. The sample values lie within the bootstrapped confidence intervals and the bootstrap mean values are very well-aligned with the sample values, thus indicating accurate estimations.

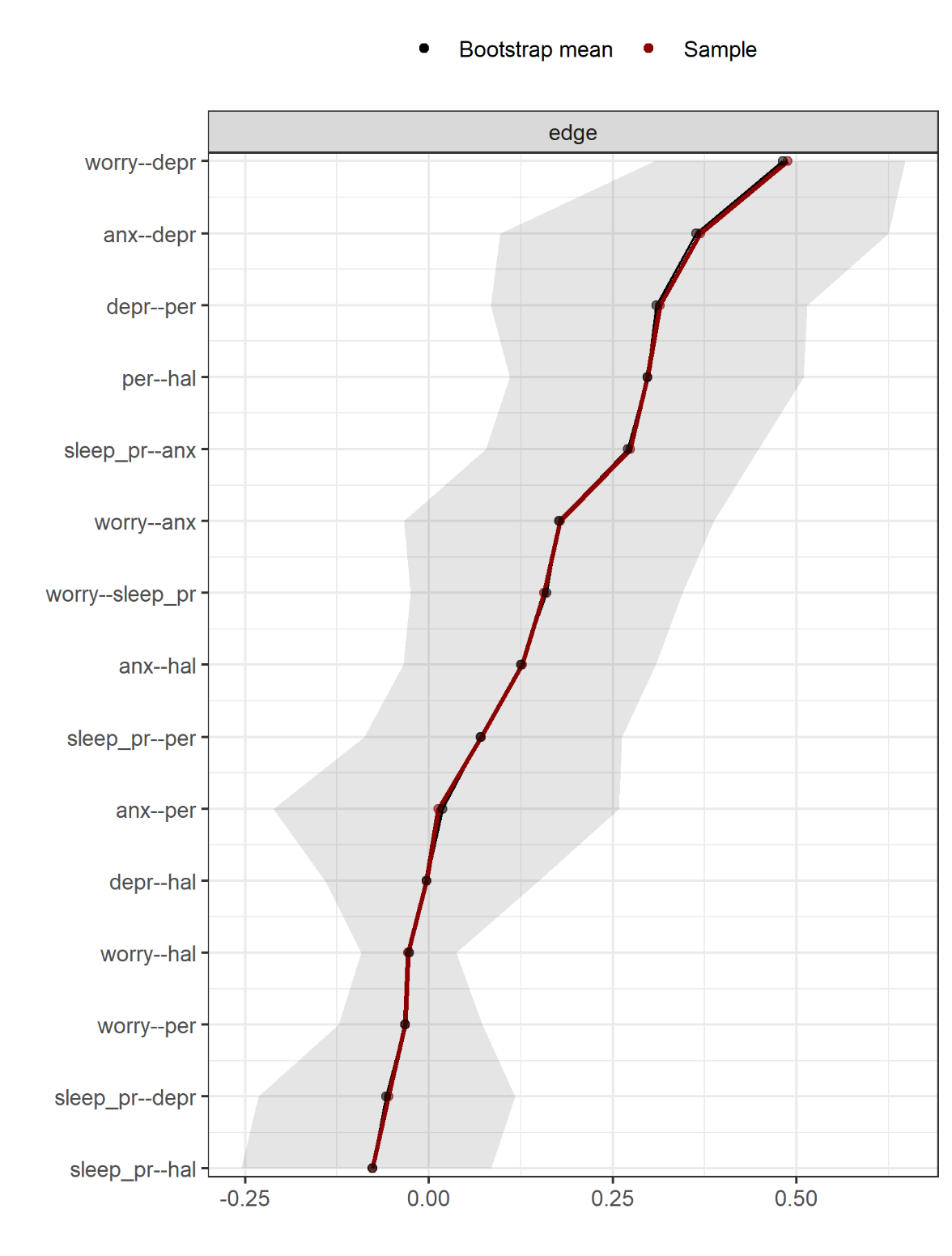

**Supplementary figure 16.** Accuracy of the edge-weights in the network estimated in men who reported neither domestic violence nor cannabis use and reported having a Black or South Asian ethnic background. The grey horizonal area within the plot represents the 95% quantile range of the parameter values across 5000 bootstraps. The red dots indicate the sample values for the analyzed data, while the black dots indicate the bootstrap mean values. The sample values lie within the bootstrapped confidence intervals and the bootstrap mean values are well-aligned with the sample values, thus indicating accurate estimations. Of note, the bootstrapped confidence intervals are relatively wide, thus some caution is recommended especially when interpreting the strength of weaker edges.
